# Next-generation biomonitoring in a cohort of pregnant women from rural Bangladesh

**DOI:** 10.1101/2025.07.22.25331978

**Authors:** Max L. Feuerstein, Md Zakir Hossain, Nicholas N.A. Kyei, Sabine Gabrysch, Benedikt Warth

## Abstract

The exposome represents all chemical exposures individuals encounter throughout their lifetime. Exposure to harmful chemicals during early life can lead to adverse later-life health outcomes, with prenatal exposure being of particular relevance. Maternal urine samples represent a valuable resource for monitoring the exposome during pregnancy including exposures to environmental, food-, and lifestyle-related toxicants. We examined 446 urine samples from pregnant women living in rural Habiganj district in Bangladesh who participated in the Maternal Exposure to Mycotoxins and Adverse Pregnancy Outcomes (MEMAPO) cohort study. Using a targeted multi-class next-generation human biomonitoring (HBM) LC-MS/MS assay, we analyzed more than 100 xenobiotics. In total, 62 target compounds were detected in urine samples, showing varying individual exposure patterns. Mycoestrogens were detected in one third of the samples, antibiotics were present in almost two thirds of the samples, and most samples contained biomarkers of exposure to polyaromatic hydrocarbons. Personal care product-related compounds, phytoestrogens, and biomarkers of exposure to nicotine, pesticides, plasticizers, and industrial chemicals were ubiquitously detected. Correlations between analytes revealed associations among chemically or functionally related compounds. This is, to the best of our knowledge, the most comprehensive HBM dataset for pregnant women in Bangladesh and South Asia. The presented dataset includes normalized urinary concentrations based on creatinine ratios and specific density. Together with previously published mycotoxin exposure data in this cohort, the presented dataset may serve as a basis for future investigations of how the detected chemical exposure mix affects pregnancy outcomes.

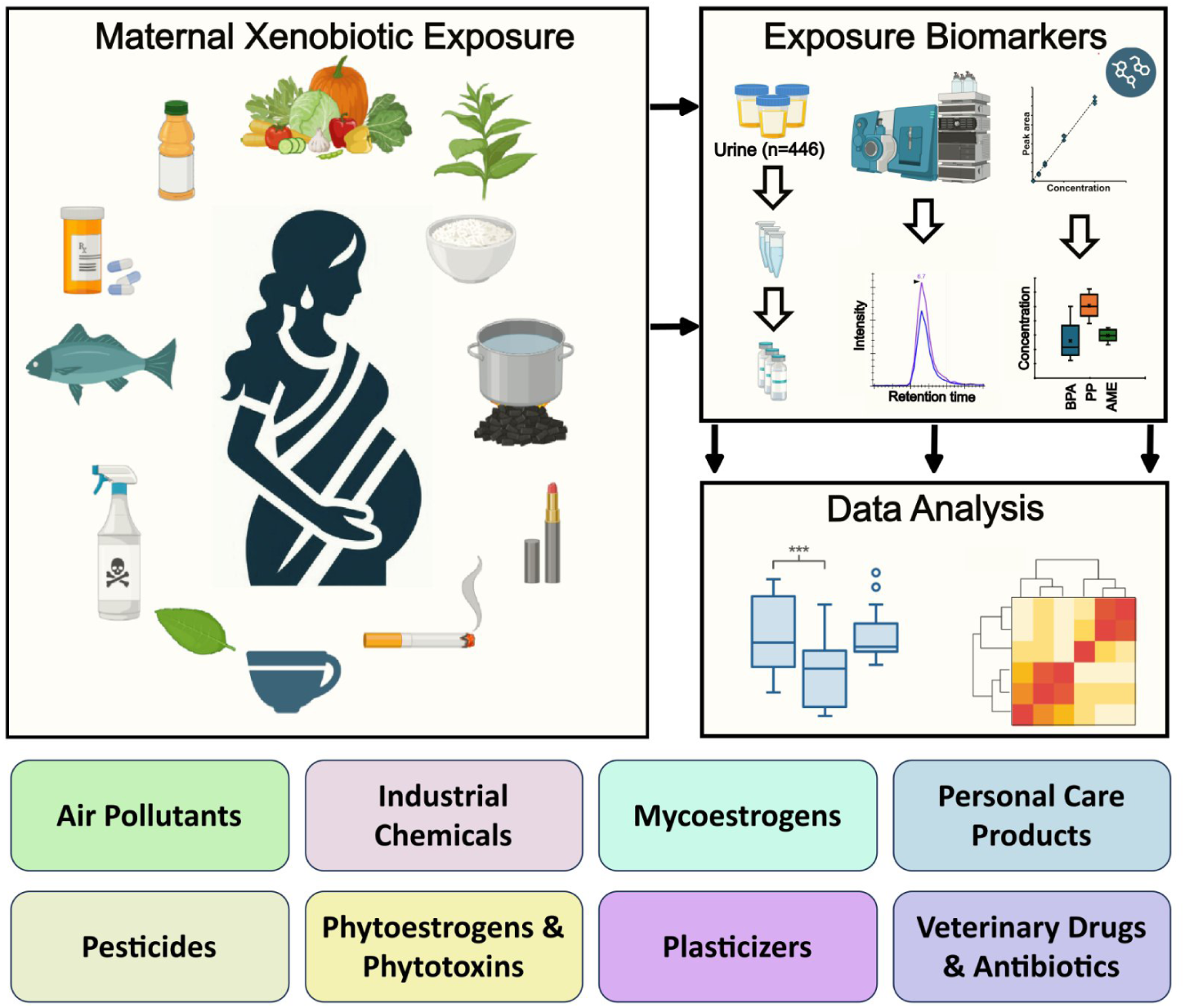

## Introduction

Environmental pollution is a major global threat, and suitable biomarkers are needed to assess the exposure and effect of harmful chemicals, which can have substantial consequences for human health. First conceptualized two decades ago, exposomics seeks to address environmental exposures as comprehensively as current technology allows (Wild, 2005, 2012; Miller, 2024). Unintended exposure to xenobiotics during early life can impact development, for example, by disrupting the endocrine system or affecting the gut microbiome, and may have adverse effects on health later in life (Ayeni et al., 2022; Krausová et al., 2023). Human exposure to environmental and food-related pollutants and contaminants can be assessed by analyzing food and environmental samples in combination with questionnaire data or by direct measurement of biological samples. Human biomonitoring (HBM) studies aim to directly address the internal concentrations (i.e., the actual body burden) of diverse toxins, e.g. in urine serum or plasma samples. Among these, urine is particularly attractive for large-scale HBM studies due to high concentrations of various urinary biomarkers of exposure, and the ease of collection – sampling is non-invasive and can be done by participants themselves (Apel et al., 2023; Esteban and Castaño, 2009). As such, HBM offers an essential toolbox for examining the effects of chemical exposure and identifying potential exposure routes, especially in cases of limited awareness of exposure or exposure sources. Traditionally, HBM studies are performed using tailored assays focusing on single compounds or compound classes, such as mycotoxins (Krausová et al., 2024; Kyei et al., 2022), parabens (Pacyga et al., 2022), or pesticides (Garí et al., 2018; Jaacks et al., 2019). Effectively addressing the complexity of the chemical exposome, however, requires highly sensitive multi-analyte assays. In recent years, several comprehensive next-generation HBM methods to assess the human exposome with broader coverage have been developed and analytically validated (Gu et al., 2025; Hossain et al., 2024; Jagani et al., 2022; Jamnik et al., 2022). Additionally, large-scale HBM programs targeting prioritized substances became well-established in regions such as Europe and the USA (Calafat, 2012; Gilles et al., 2021). Due to its excellent sensitivity, specificity, and robustness, tandem mass spectrometry, coupled with high-performance liquid chromatography (LC-MS/MS), is the most frequently used analytical technology for large-scale HBM studies. Extending analyte panels for such HBM assays is a current priority to ensure broader, exposome-scale coverage (Hossain et al., 2024). For instance, antibiotics are not routinely monitored in large HBM studies, despite growing evidence of exposure to residual antibiotics and related adverse health effects (Hossain et al., 2025; Hu et al., 2022; Zhu et al., 2020). While broad HBM and exposome-oriented cohort studies are becoming more routine, most large-scale studies focus on economically developed world regions and countries (Calafat, 2012; Gilles et al., 2021). In less affluent world regions, HBM data are often scarce and limited to small sets of environmental pollutants, occupational exposures, and food contaminants (Ayeni et al., 2022; Barnett-Itzhaki et al., 2018). Thus, more efforts are required to close research gaps on environmental health risks in potentially highly exposed populations in low- and middle-income countries, including those related to emerging contaminants such as plasticizers (Acevedo et al., 2025; de Paula Nunes et al., 2025).

In Bangladesh, widespread pollution with heavy metals, arsenic, pesticides, and residual antibiotics has been reported in ground and surface water as well as in drinking water (Angeles et al., 2020; Chen et al., 2011; Kibria et al., 2016; Parvin et al., 2022). Air pollution originating from traffic and biomass combustion (Barnett-Itzhaki et al., 2018), agricultural pesticide contamination (Khatun et al., 2023), and exposure to mycotoxins (Kyei et al., 2022) pose significant risks to human health. Yet, to the best of our knowledge, only a limited number of comprehensive HBM studies have been conducted to date, mainly focusing on single analyte classes like organochlorines (Bergkvist et al., 2012), heavy metals (Forsyth et al., 2019), arsenic (Chen et al., 2011), pesticides (Jaacks et al., 2019), or mycotoxins (Kyei et al., 2022). Building on these studies, we applied a recently scaled-up multi-class HBM method for analyzing over 100 target compounds to comprehensively investigate co-exposure patterns in a cohort in rural Bangladesh, allowing a detailed assessment of the chemical exposome of pregnant women from rural Bangladesh for the first time. This study also represents the first large-scale application of a recently validated HBM assay (Hossain et al., 2024). Diverse classes of natural and synthetic chemicals, including personal care products, pesticides, plasticizers, and phytoestrogens, were investigated, including various endocrine-disrupting chemicals. Additionally, a set of endogenous estrogens and their metabolites was included in the analyte panel (Hossain et al., 2024; Preindl et al., 2019), enabling the simultaneous investigation of chemical exposure and the potential effects of total xenoestrogenic burden on hormone levels using a single analytical assay.

## Materials and Methods

### Study population

The *Food and Agricultural Approaches to Reducing Malnutrition* (FAARM) cluster-randomized controlled trial was conducted in two rural sub-districts of Habiganj district, Sylhet division, Bangladesh (ClinicalTrials.gov ID: NCT02505711). The trial enrolled 2705 married women from 96 settlements who reported being 30 years or younger, having access to at least 40 m² of land and declared an interest in gardening (Wendt et al., 2019). As an extension of FAARM, the *Maternal Exposure to Mycotoxins and Adverse Pregnancy Outcomes* (MEMAPO) cohort study followed 439 pregnant FAARM participants from early pregnancy to investigate the effects of maternal mycotoxin exposure on adverse pregnancy outcomes, as well as associations with sociodemographic characteristics and consumption of specific foods and local stimulants (Kyei et al., 2023, 2022), further details provided in the **Supplementary Information**. Mycotoxin exposure in this cohort was substantial, with high detection frequencies in urine for ochratoxin A (95%) and citrinin (61%), and higher concentrations in samples from women who had consumed chewing tobacco or betel nut/leaf in the past month (Kyei et al., 2022). Moreover, exposure to ochratoxin A was associated with an increased risk of having a low birth weight baby (Kyei et al., 2023).

### Urine sample collection and processing

We analyzed 446 urine samples of 438 pregnant women (some were pregnant twice, one sample was lost) who consented to participate in the MEMAPO study in Bangladesh and provided a urine sample before 20 weeks of gestation. Between July 2018 and November 2019, 10 mL of first-morning urine was collected in a sterile disposable container by study participants before consuming food and water.

Samples were transported to the project field laboratory in a cold box, aliquoted into 2 ml safe-seal tubes, and stored at −20°C on the same day. Aliquots were intended for the analysis of mycotoxins and other environmental chemicals. Subsequently, all samples were sent to Germany on dry ice (transport time < 24 h) and stored at −70°C at Heidelberg University Hospital. Sample aliquots for chemical exposome analysis were later transported on dry ice to the University of Vienna, Austria (transport time <6 h) and stored at −80°C until biomarker analysis. The sociodemographic characteristics of the women providing the urine samples are summarized in **Table S1**. The findings on mycotoxin exposure as part of MEMAPO have been published previously (Kyei et al., 2023, 2022).

### Chemicals and reagents

LC-MS grade water, methanol (MeOH), and acetonitrile (ACN) were obtained from Honeywell (Riedel-del Haen, Germany) and VWR chemicals (USA). Ammonium fluoride (NH_4_F) was purchased from Honeywell (Fluka, Germany), ammonium acetate (NH_4_Ac), and acetic acid from Sigma-Aldrich (Darmstadt, Germany). A total of 136 authentic reference standards from diverse chemicals and a mix of isotopically labeled internal standards (ISTDs) were purchased from Sigma Aldrich and Toronto Research Chemicals (Canada). A complete list of used standards and ISTDs can be reviewed elsewhere (Hossain et al., 2024; Jamnik et al., 2022) and as part of the **Supplementary Information**. Stock solutions of individual analytical standards were dissolved in LC-MS grade acetonitrile, methanol, water, or dimethyl sulfoxide at 1 mg/mL or 2 mg/mL. ISTDs were prepared from individual stock solutions (1 mg/mL in ACN or MeOH). A working stock solution containing analytical standards was prepared in ACN, and calibration standards were prepared at eight different concentration levels. For internal quality control, the ISTD mix was spiked into the samples before sample extraction. All authentic standards and stock solutions were stored at −20°C until the day of use. Complete chemical information, including all concentration levels of calibration standards and ISTDs, is provided in **Tables S2-4**. A summary of all abbreviations used to refer to target analytes is included in **Tables S2** and **S6**.

### Sample extraction

A protein precipitation sample preparation protocol was used for the extraction of analytes from urine (Hossain et al., 2024; Preindl et al., 2019). All samples, standards, and solutions were kept on ice between sample preparation steps to avoid analyte degradation. In brief, 100 μL of urine was mixed with 400 μL of ACN/MeOH (1/1, v/v) containing the ISTD mixture. Samples were vortexed and sonicated for 10 min in an ice bath, and proteins were precipitated for 2 h at −20°C. Samples were then centrifuged in a pre-cooled centrifuge (4°C, 10 min, 18,000×g), and the supernatants were collected and dried in a CentriVap vacuum concentrator (Labconco) at 7°C. After evaporation, samples were reconstituted in 100 μL ACN/H_2_O (10/90, v/v), vortexed, and centrifuged (4°C, 10 min, 18,000×g). Supernatants were transferred to glass vials with inserts and were stored at −20°C until the day of measurement. Finally, samples were thawed and vortexed before LC-MS/MS measurements. In addition, enzymatic deconjugation (i.e., β-glucuronidase/sulfatase treatment) was performed for a subset of ten samples using one of our previously published protocols (Fareed et al., 2022; Hossain et al., 2024). In short, 100 µL of urine was treated with 100 µM β-glucuronidase/sulfatase enzyme solution from *Helix pomatia* in 2.5 M NH_4_Ac buffer adjusted to pH 5.5. Samples were incubated for 16 h at 400 rpm and 37°C on a shaker followed by extraction using the same protocol as described for non-treated urine samples and using a final reconstitution volume of 100 µL. The resulting dataset was used as the basis to test the feasibility of the used approach. Estimated ratios of free and total analyte concentrations were then compared to literature values where applicable. The full sample set was intentionally not treated with β-glucuronidase/arylsulfatase enzymes. This was done to (1) allow for potential re-analysis of these extracts from finite high-value samples for advanced non-targeted analysis and suspect screening (NTA/NTS) by high-resolution mass spectrometry (Guerrini et al., 2024; Lai et al., 2024) and (2) because a recent systematic evaluation demonstrated that these enzyme mixtures are often highly contaminated by a vast number of xenobiotics, including many of the target analytes of the utilized assay (Fareed et al., 2022). In the presented work, we reproduced some observations and detected notable contaminations of the enzyme mix for a diverse set of natural and synthetic toxins (details presented in the **Supplementary Information**).

### LC-MS/MS data acquisition

LC-MS/MS was performed using a recently expanded and in-house validated method for the analysis of various classes of exposure chemicals (Hossain et al., 2024). All samples were analyzed using a 1290 Infinity II UHPLC system (Agilent) coupled to a Sciex QTrap 6500+ mass spectrometer (Darmstadt, Germany) equipped with a Turbo-V^TM^ electrospray ionization (ESI) source operating in fast polarity switching mode. A reversed-phase Acquity HSS T3 column (1.8 μm, 2.1×100 mm) equipped with a VanGuard (1.8 μm) pre-column from Waters (Vienna, Austria) was used for front-end separation. Autosampler and column oven temperatures were set to 7°C and 40°C, respectively, and an injection volume of 5 µL was used. Chromatographic gradient separation was performed using an aqueous eluent (water containing 0.3 mM of ammonium fluoride) and an organic eluent (98% ACN and 2% water containing 0.3 mM of ammonium fluoride) at a flow rate of 0.4 mL/min and a runtime of 20 min. The detailed chromatographic gradient has been described elsewhere (Hossain et al., 2024). The SCIEX 6500+ QTrap was controlled by the Analyst software (version 1.5.5), and a scheduled multiple reaction monitoring (sMRM) method was used for data acquisition. The MS was operated using the following parameters: ion spray voltage was −4.5 kV in negative mode and 5.5 kV in positive mode. Nitrogen was used as the ESI gas and as the collision gas. The collision gas (CAD) pressure was set to “medium”, curtain gas was set to 30 psi; ion source gas 1 was 80 psi, and ion source gas 2 was 60 psi. Additional detailed information on MS and MS/MS parameters, including polarity, Q1, Q3, DP, CE, and CXP, retention times, and monitored transitions, has been provided elsewhere (Hossain et al., 2024; Jamnik et al., 2022).

### Batch design and quality control strategy

Analysis was performed in a blinded manner and in batches of 20-30 samples. During the measurement of experimental samples, different types of QC-samples were analyzed: (1) solvent blanks were injected at the start of the sequence, before and after solvent calibration, and after 20-30 urine samples to track potential carryover, and (2) three process blanks were injected as triplicates to determine background contaminations from the sample preparation process. In addition, (3) a pooled urine sample, donated by a female volunteer who reduced exposure to xenoestrogen by avoiding foods or cosmetics stored in plastic containers and foods rich in phytoestrogens for two days before sample collection (Preindl et al., 2019), was used to prepare quality control samples. In addition to non-spiked pooled urine QCs, three replicates of (4) spiked urine samples were included. For this purpose, analytical standards were spiked into the QC sample before analyte extraction, using a concentration corresponding to the calibration standard at “level 30” (see **Table S3**). **Figure S3** shows example chromatograms and peak areas (Area under curve; AUC), illustrating the repeatability of the assay over the entire sequence (i.e., 700 injections). Three replicates of the non-spiked and spiked QC samples were extracted and measured at the beginning and the end of the sequence to estimate the apparent recovery. Moreover, spiked quality control samples were measured at the beginning of every analytical batch to monitor potential shifts in retention time or detector sensitivity. (5) ISTDs were added to every sample at the beginning of the extraction process as an internal quality control. Ten selected ISTDs were monitored throughout the entire analytical sequence to ensure consistent retention times, extraction, and instrument response. Data for the ISTDs are presented in **Table S5** and **Figure S1.** Finally, two aliquots of urine samples from two individuals were analyzed as (6) blinded QC samples to monitor the consistency of replicate extraction and analysis of real-life samples. Only analytes with sufficient method linearity (i.e., with R^2^>0.9), extraction recovery, and stability of retention times over the entire analytical sequence are reported herein.

### Data processing and data analysis

Data processing was performed using SCIEX OS 3.0 and Skyline (McCoss Lab) (Adams et al., 2020) for peak picking, data extraction, peak review, and analyte quantification. In SCIEX OS 3.0 the “Autopeak” integration algorithm was used. Integration parameters were optimized for each analyte, false positives were curated through expert review, and integration boundaries were manually adjusted as needed. Peaks with signal-to-noise ratios (S/N) <3 were manually removed. A weighted linear regression model (1/x) was used to build calibration curves from calibration standards in solvent. Weighted linear regression models were selected to ensure comparability to the originally published methods and to ensure good quantification accuracy for low-abundant analytes. Analytes were quantified based on peak areas, and internal standards were used to track the repeatability of extraction and analysis. Slope, intercept, and R^2^ were used to test linearity and are provided in **Table S6.** MRM-chromatograms were investigated, and compounds with strong retention time shifts, interfering peaks, poor linearity (i.e., calibration curves with R^2^<0.9), or compounds that couldn’t be recovered from spiked QC samples were not reported. Results tables were exported, and further calculations, including blank correction and correction for apparent recovery, were performed using R together with R-Studio or using Microsoft Office (Professional Plus 2019). Blank correction was performed by subtracting the average concentration of three extraction blanks, and apparent recoveries were used to correct for matrix effects and analyte recovery (details presented in the **Supplementary Information**). Reported data were classified as quantitative for analytes with apparent recoveries between 60% and 140%, whereas results for compounds outside these specifications are reported as semiquantitative (**Table S6**). For full details of the method validation, including limits of detection (LODs) and limits of quantification (LOQs), we refer the reader to our previous work (Hossain et al., 2024; Jamnik et al., 2022). Detection frequencies were calculated as fractions of samples with analyte concentration >LOD. For further calculations, concentrations <LOD were replaced with 0.5·LOD, and concentrations <LOQ were replaced with 0.5·LOQ, and mean, median, and 95^th^ percentile concentrations were calculated for all samples after data imputation (**Table S6**). The resulting data were then compared with literature, reference concentrations, and human biomonitoring guidance values (HBM-GVs) where available (**Tables S7** and **S8**). For this purpose, the ratios of free and total urinary biomarker concentrations were estimated using literature values and experimental estimates determined during this study, with details presented in the **Supplementary Information**. Moreover, urinary concentrations, as listed in **Table S9**, were normalized to specific gravity and urinary creatinine concentrations (see **Tables S10-S12**). Two urine samples were included as duplicates and were measured as a blinded QC to ensure suitable repeatability during the sequence (see **Table S13**). Spearman correlation coefficients were determined to investigate relationships between detected xenobiotics and endogenous estrogen levels. Skyline (v23.1) was used to inspect data and create figures. R, together with RStudio, was used to plot results and conduct further data analysis, including Spearman correlation. Inkscape, Microsoft Office, and Biorender were used to create graphics and edit figures. The graphical abstract was created in BioRender (Feuerstein, M. (2025) https://BioRender.com/pl2mq25).

### Assessment of HBM guidance values

Performing a full risk assessment, including toxicokinetic models and consideration of potential mixture effects, was outside the scope of this work and would require further mechanistic studies. However, to assess potential risks related to chemical exposure, urinary concentrations were compared with established HBM-GVs, HBM-I values, or population-based reference values, where available. Referenced data are presented in more detail in **Table S7** and **Table S8**. Notably, the majority of reference values for urinary biomarkers of exposure are based on total urinary concentrations after deconjugation, which limits comparability with the data presented herein. To allow comparisons across datasets and with established HBM-GVs, estimates for the ratio of free and total concentrations derived from experimental data or published literature were applied (see **Table S15**).

## Results and Discussion

### Quantification of xenobiotics and estrogens in urine samples

Taking advantage of a recently developed, sensitive LC-MS/MS method, a large number of diverse exposure chemicals could be investigated simultaneously in urine. In total, 97 targeted analytes were recovered during the extraction of spiked QC samples, had sufficient method linearity (calibration curve with R^2^>0.9), stable retention times over the complete analytical sequence, no interfering signals, and no carryover. The remaining compounds were classified as “not reported” in **Table S6** to ensure high data quality. **Figures S4** and **S5** show example chromatograms for detected compounds in urine samples. A total of 62 analytes were detected in at least one sample. These included air pollutants, industrial chemicals, mycotoxins, personal care products, pesticides and pesticide metabolites, phytoestrogens, phytotoxins, plasticizers and plastic-related compounds, and antibiotics. Analyte concentrations were highly diverse, spanning from a few pg/mL to >1000 ng/mL. The following xenobiotics were present with detection frequencies (DF) >75%: the personal care product-related parabens methylparaben and propylparaben, the industrial chemical 2-naphthol, the nicotine metabolites cotinine and trans-3-OH-cotinine, the plasticizer n-butylbenzensulfonamide, the phytoestrogens daidzein, genistein, enterolactone, and enterodiol, and the pesticide metabolites p-nitrophenol and diethyl thiophosphate (DETP). Other potentially harmful chemicals were detected in substantial fractions of the samples. For instance, the plastic-related chemicals bisphenol A (BPA), the mycotoxin alternariol monomethyl ether (AME), and the pesticide imidacloprid (IDC) were detected in more than 25% of all samples. **Figure 1** summarizes the detection frequencies and concentrations of the targeted analyte panel in urine samples. Example MRM-chromatograms for positive detections and blank samples are presented alongside the peaks in spiked QC samples for the pesticide imidacloprid (IDC) and the mycotoxin alternariol (AOH). In addition to relevant xenobiotics, 14 endogenous estrogens and estrogen metabolites were analyzed in the same assay, allowing for the investigation of the potential interplay between exposure to xenoestrogens and endogenous estrogen concentration. In total, ten endogenous estrogens were detected in urine: estrone (E1; DF = 99%, concentration range: <LOQ −240 ng/mL), estradiol (E2; DF = 86%, range: <LOQ −54 ng/mL) and estriol (E3; DF = 96%, range: <LOQ −500 ng/mL) were detected in most analyzed urine samples, whereas several other estrogen metabolites were found with lower detection frequencies and with lower urinary concentrations. For example, 4-methoxy estrone (4MeOE1) was detected in 25% of the samples with concentrations ranging from <LOQ to 0.092 ng/mL. A detailed overview of the results, including detection frequencies, mean, median, minimum, maximum, and 95^th^ percentile concentrations, is presented in **Table S6**.

**Figure 1:**
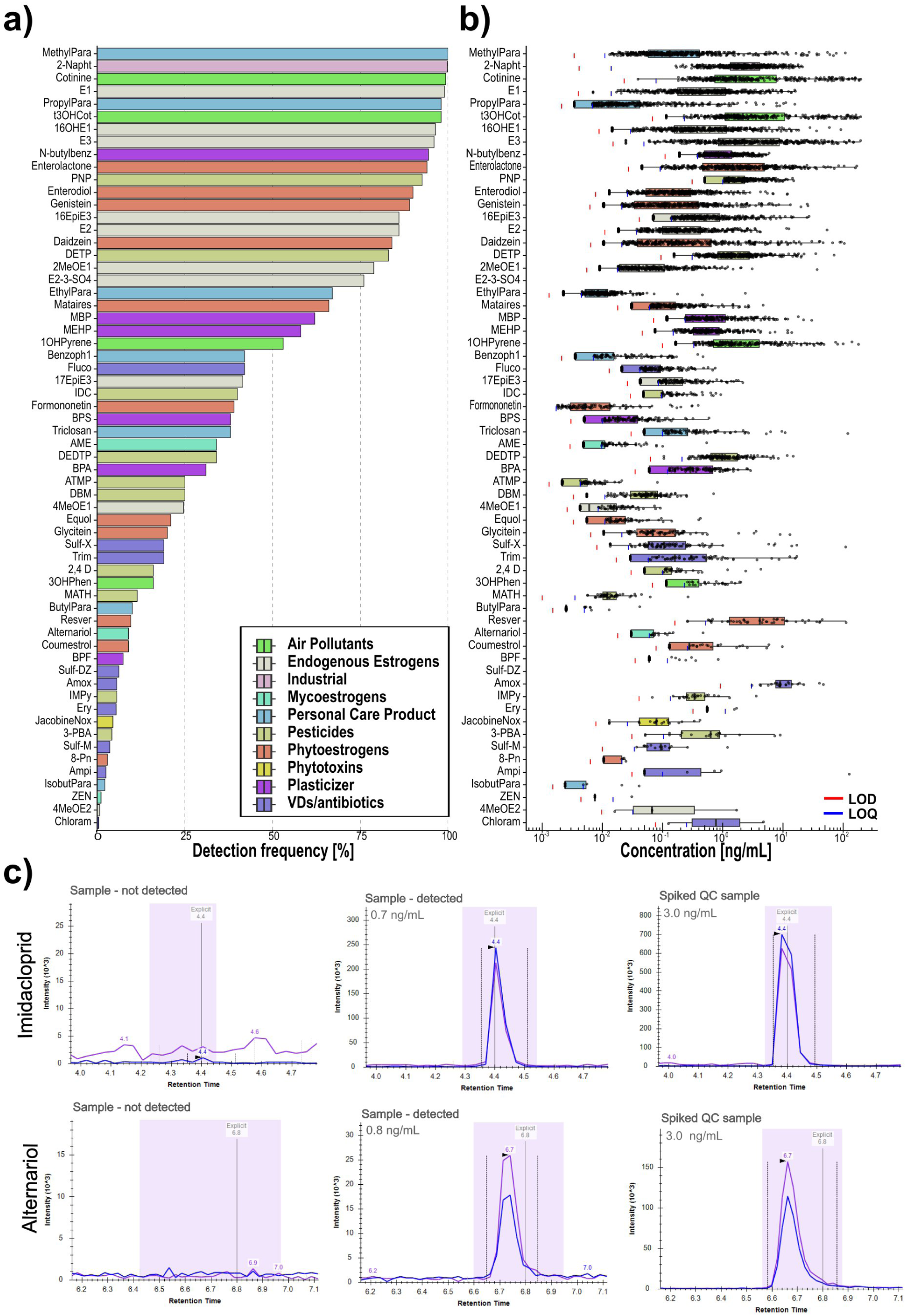
a) Detection frequencies of detected analytes (n=62) in the urine samples (n=446), b) boxplots showing urinary concentrations and c) extracted ion chromatograms (EICs) for the pesticide imidacloprid (IDC), and the mycotoxin alternariol. From left to right panels show examples for blank samples (i.e., analyte not detected), detected compounds in samples and a spiked quality control (QC). For a full list of abbreviations, please refer to **Table S6**. Only concentrations >LOD are shown and concentrations <LOQ were imputed with 0.5·LOQ for the boxplots.

### Deconjugation and estimation of free total concentrations of targeted analytes

Typically, HBM assays focus on a few classes of analytes per assay, and in the case of urine analysis, tailored enzymatic deconjugation is widely used. However, we recently showed that, for broad-scale HBM, optimizing enzymatic deconjugation protocols is challenging and often requires balancing enzymatic efficiency against the risk of process-related contaminations (Fareed et al., 2022). Nevertheless, measuring total concentrations rather than the free fraction is often required for the analysis of urine samples to ensure comparability to established reference concentrations and HBM-GVs. To improve the comparability of our data, we measured a subset of 10 urine samples after enzymatic treatment and compared the ratios of free to total concentrations of selected analytes to literature values, where available (see **Table S15**). Results after performing deconjugation with *H. pomatia* are presented in detail in **Table S14** and **Figure 2**. Notably, relevant background contamination was introduced during sample treatment with *H. pomatia* extracts, and 27 analytes were detected in procedural blanks after enzymatic treatment. These compounds included various natural and synthetic toxins such as alternariol, alternariol monomethylether (AME), 2-naphthol, and several phytoestrogenic compounds. Moreover, the mycotoxin zearalenone (ZEN), the pesticide atrazine, and the toxic alkaloid aristolactam were detected, resulting in limited applicability of the workflow for these analytes (see **Table S16** and **Figure S8**). Nevertheless, measuring total concentrations clearly increased the assay sensitivity for several analytes, including the PAH metabolites 1-hydroxypyrene, 3-hydroxyphenanthrene, monobutylphthtalate (MBP), monoethylhexylphthalate (MEHP), ethylparaben and propylparaben, bisphenol-A (BPA), triclosan, and several endogenous estrogens. Reporting only the free (i.e., non-conjugated) analyte form results is an underestimation of the actual urinary concentrations for these analytes. For instance, MBP was detected in 9/10 of the samples after deconjugation, whereas the free form was only detected in 4/10 of the samples. Noteworthy, only 10 samples were analyzed after deconjugation and low detection frequencies of the free parent analytes allow only a rough estimate of the relative contribution of the free form to total urinary concentrations for many analytes. However, these estimates were in general agreement with literature values, with free analyte concentrations accounting for approximately <1% to 10% of total urinary concentrations for most analytes. Thus, literature values were used (when available) to estimate the free-to-total concentration ratio for several exposures. In the absence of suitable reference values, experimental estimates were used. For instance, the free fraction accounted for <1% of the total concentration for enterodiol and enterolactone.

**Figure 2:**
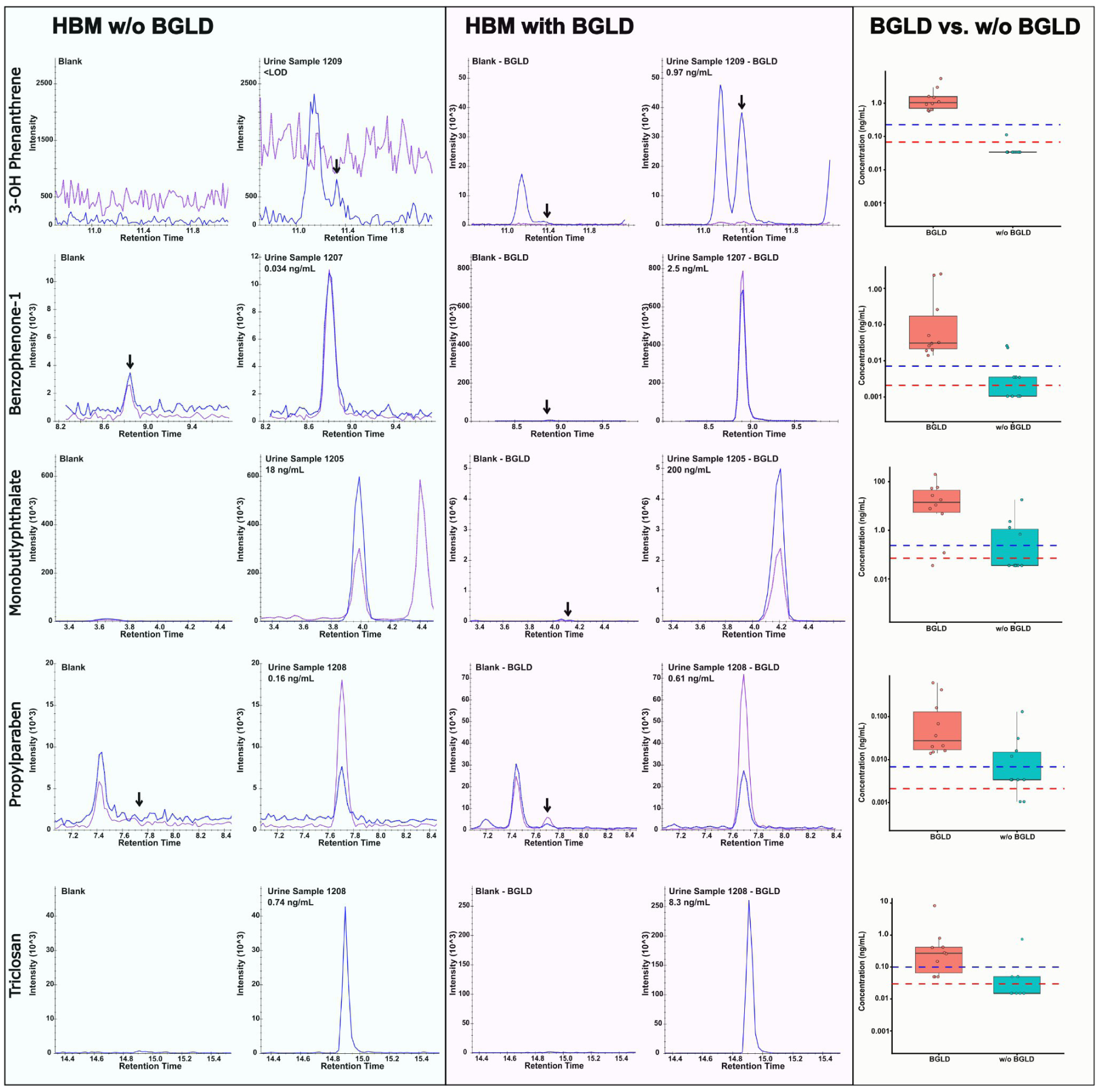
multiple reaction monitoring (MRM)-chromatograms of extraction blanks, deconjugated samples using β-glucuronidase/sulfatase from *Helix pomatia* (BGLD), and samples without using enzymatic deconjugation (w/o BGLD). Boxplots show the corresponding concentrations for a subset of ten urine samples. Data are presented for 3-hydroxy phenanthrene, benzophenone-1, monobutlyphthalate, propylparaben and triclosan. Limits of detection and quantification are represented as red and blue dashed lines. Concentrations <LOQ were replaced with 0.5·LOQ and <LOD with 0.5·LOD in the plot.

### Air pollutants and nicotine metabolites

Four compounds classified as potential metabolites of air pollutants were detected in urine: The polyaromatic hydrocarbon (PAH) metabolites 1-OH-Pyrene (1OHPyr) and 3-OH-phenanthrene (3OHPhen), and the two nicotine metabolites cotinine and trans-3-hydroxycotinine (t3OHCot) (see **Figure 1**). 1OHPyr and 3OHPhen were detected in 53% and 16% of all samples, respectively, with concentrations between <LOQ −180 ng/mL and <LOQ −2.0 ng/mL. These analytes are human metabolites of the PAHs phenanthrene and pyrene and are markers for exposure to air pollution (e.g., due to traffic or tobacco smoke), biomass combustion (e.g., cooking on open fire (Barnett-Itzhaki et al., 2018)), consumption of smoked and barbecued food (Murawski et al., 2020), or other consumer goods, including diverse tobacco products (McAdam et al., 2013).

Although only free fractions were measured, the concentrations and detection frequencies were higher than in a recent study from Slovenia (Joksić et al., 2022), where 1OHPyr was detected in 14% of all urine samples with 95^th^ percentile concentration of 0.17 ng/mL, and 3OH-phenanthrene in 10% of samples with 95^th^ percentile concentration of 0.16 ng/mL (see **Table S8**). Similar observations were made when comparing the results of this study with the 95th percentile reference value (RV95) for female adolescents in Germany (Hoopmann et al., 2023; Murawski et al., 2020) (see **Table S7**). Notably, both datasets are based on enzymatic deconjugation during sample preparation. In contrast, only free analyte fractions were determined in the work at hand, which makes a direct comparison of the data challenging. Based on deconjugation of 10 samples, we estimated that the free forms in urine account for <1% and <3% of the total 1OHPyr and 3OHPhen, respectively. Given this high degree of conjugation, most samples tested positive for free 1OHPyr and 3OHPhen, exceeding reference values and indicating a generally high exposure to PAHs in the studied population, which is in line with the widespread use of solid fuels for cooking in rural Bangladesh (Rasel et al., 2024).

Nicotine metabolites cotinine and t3OHCot were detected in 99% and 98% of samples, respectively, with concentrations up to 1500 ng/mL and 5800 ng/mL, respectively. Both compounds are typically found in high concentrations in individuals exposed to tobacco smoke (i.e., via active or passive smoking), but could also result from other forms of nicotine intake. Due to the unavailability of more suitable reference values, urinary concentrations in this study were compared to RV95 concentrations for female adolescents from Germany (Hoopmann et al., 2023) (see **Table S7**). Urinary cotinine concentrations exceeded the RV95 value in 44% of cases among non-smoking adolescents. Thus, our findings align with the widespread use of chewing tobacco (*jorda*) and other local stimulants in the studied cohort. Two months before urine sampling, 43% of the women reported use of stimulants during the previous 4 weeks (Kyei et al., 2022).

### Industrial chemicals

Nonylphenol was detected in the majority of samples, yet due to relatively high procedural blanks, the data are not reported. In future studies, additional biomarkers for nonylphenol exposure, such as its main urinary biomarker hydroxynonylphenol, may serve as indicators of nonylphenol exposure (Ringbeck et al., 2021). The PAH metabolite 2-naphthol (DF = 100%, range: 0.17–21 ng/mL) was ubiquitously detected in urine samples. As a primary metabolite of naphthalene, 2-naphthol serves as a biomarker for exposure to PAHs, which can originate from sources such as traffic emissions, tobacco smoke, and industrial processes. 2-naphthol is commonly detected in HBM studies (Hartmann et al., 2023; Jamnik et al., 2022; Joksić et al., 2022). For instance, a recent study from Slovenia detected 2-naphthol in 100% of the investigated urine samples from lactating women. Although this study used deconjugation during sample preparation, slightly lower urinary concentrations were observed compared with our study (95th percentile = 8.3 ng/mL, compared with 11 ng/mL in urine samples from Bangladesh) (Joksić et al., 2022). Moreover, RV95 values for German adolescents of 22 ng/mL for total 2-naphthol in urine were not exceeded when only the free form was considered in our study (Hoopmann et al., 2023). Due to the high concentrations in procedural blanks (33 ± 1 ng/mL) after deconjugation using glucuronidase and sulfatase enzymes from *H.pomatia*, an experimental estimation of free and total concentration was not feasible. Yet, similar to other PAH-metabolites, 2-naphthol is mainly excreted in conjugated forms in urine, and the free form only accounts for a few percent of the total concentration. For instance, Gaudreau et al. reported 4% free 2-naphthol in urine samples (Gaudreau et al., 2016). Using this as an estimate for the ratio of free and total urinary 2-naphthol concentrations, approximately 86% of all analyzed samples exceed RV95 for adolescents from Germany, and 98% exceed 95^th^ percentile urinary concentrations in urine samples of lactating women from Slovenia (Hoopmann et al., 2023; Joksić et al., 2022).

### Antibiotics and veterinary drug residues

In total, nine out of 17 analyzed antibiotics were detected in at least one urine sample, with the highest detection frequencies reported for fluconazole (DF = 42%, range: <LOQ −0.75 ng/mL), trimethoprim (DF = 19%, range: <LOQ −5000 ng/mL), and sulfamethoxazole (DF = 19%, range: <LOQ −110 ng/mL). Additionally, sulfadiazine (DF = 6.2%, range: <LOQ −6.4 ng/mL), amoxicillin (DF = 5.6%, range: 3.1 ng/mL −39000 ng/mL), erythromycin (DF = 5.4%, range: <LOQ −1.7 ng/mL), sulfamethazine (DF = 3.6%, range: 0.037 −1.2 ng/mL), ampicillin (DF = 2.5%, range: <LOQ −130 ng/mL), and chloramphenicol (DF = 0.5%, range: <LOQ −4.7 ng/mL) were detected (see, **Table S6**). In total, 64% of the analyzed urine samples contained at least one of the 17 targeted antibiotics and up to five antibiotics were detected per urine sample (see **Figure S6**). Most detected antibiotics can be used to treat humans and animals alike, and exposure routes remain unclear, without detailed questionnaire data or medical records on antibiotic intake. High concentrations in a small number of samples, e.g., as observed for trimethoprim or amoxicillin, can be regarded as an indication for active intake due to medical treatment, whereas high detection frequencies combined with low analyte concentrations may result from other exposure routes, e.g., uptake via contaminated drinking water, fish, milk, or meat (Hu et al., 2022). Detection frequencies for several antibiotics, including trimethoprim and sulfamethoxazole, were largely comparable to those reported in a cohort from Ghana (Lerbech et al., 2014) and China (Geng et al., 2020). However, direct comparisons between datasets are constrained by variations in sample preparation methods, such as application of deconjugation or solid-phase extraction in the referenced studies (see **Table S8**). Nevertheless, frequent detection of antibiotics in human urine samples underscores the need for systematic inclusion in large-scale HBM studies (Hossain et al., 2025). Frequent detection rates may reflect widespread background exposure, which poses potential risks for antimicrobial resistance, gut dysbiosis, and related health consequences (Hu et al., 2022).

### Estrogenic mycotoxins (mycoestrogens)

Food-related exposure to mycotoxins is a global health threat, causing cancer and damaging various organs and the immune system (Wild and Gong, 2010) and may result in adverse pregnancy outcomes (Krausová et al., 2024; Kyei et al., 2020). In a previous study, frequent exposure to ochratoxin A (OTA), citrinin, and deoxynivalenol was observed in the same samples, and 95% of the samples exceeded the margin of exposure values for OTA. Urinary OTA concentrations were associated with consumption of nuts, and local stimulants such as betel nuts, betel leaves, or chewing tobacco (Kyei et al., 2022). Moreover, exposure to OTA was linked to higher odds of low birth weight (Kyei et al., 2023). To complement existing data, we focused on mycotoxins with xenoestrogenic potential, especially the *Alternaria* toxins alternariol (AOH) and alternariol monomethyl ether (AME). Although these toxins were included in the previously applied multi-mycotoxin assay, they were likely not detected due to the lower sensitivity of the previously applied method (Kyei et al., 2023, 2022). With the assay employed herein, we were able to detect AOH and AME in 8.9% and 34% of all samples, respectively, with concentrations ranging from <LOQ to 0.47 ng/mL in the case of AOH and <LOQ to 0.54 ng/mL in the case of AME. The successful detection of AOH and AME using our new multi-class exposomics assay was made possible by lower LOD and LOQ values, underscoring the importance of sufficient assay sensitivity. A recent study reported AME with comparable detection frequency in a cohort of pregnant women from the USA, utilizing a specific mycotoxin method with deconjugation (Krausová et al., 2024). Zearalenone was detected in only a small fraction of our samples at very low concentrations (DF=1.1%, range: <LOQ-0.031 ng/mL), which is consistent with previous findings for this cohort (Kyei et al., 2022). In total, 36% of all samples contained low concentrations of AOH, AME, or ZEN. Due to the high analytical background for these mycotoxins in the *H.pomatia* enzyme solution used for deconjugation, free fractions could not be estimated. Yet, frequent detection of the free fraction of AOH and AME indicates relevant exposure of the studied population.

### Personal care products

Different types of additives and preservatives are used to increase the shelf life of personal care products (PCPs) and cosmetics. Parabens represent a frequently used class of additives and preservatives in PCPs, and a wide range of endocrine-disrupting properties has been reported (Nowak et al., 2018). In this study, we detected seven PCP-related substances, with at least one PCP-related compound present in every sample. Methylparaben (DF = 100%, range: 0.013 ng/mL −670 ng/mL), propylparaben (DF = 98%, range: <LOQ −950 ng/mL), and ethylparaben (DF = 67%, range: <LOQ – 2.4 ng/mL) were detected in most samples, whereas butylparaben and isobutylparaben were only detected in 10% and 2.2% of urine samples, respectively. Additionally, the UV-filter benzophenone-1 was detected in 42% of samples (range: <LOQ −0.47 ng/mL), and the antimicrobial agent triclosan was detected in 38% of samples (range: <LOQ −120 ng/mL). The free form accounts for approximately 1-10% of total urinary concentrations and depends on the studied compound (see **Table S15**). Even after consideration of estimated free fractions of urinary PCPs, concentrations for parabens, triclosan, and benzophenone-1 were largely below the established RV95 for parabens and benzophenone-1, and the HBM-I value for triclosan in urine (**Table S7**). Only 0.2% of the samples exceeded HBM-I values for triclosan. RV95 reference concentrations established for the German population were exceeded in 0.22% for ethylparaben, 4% for proylparaben, and 2.7% for methylparaben (Apel et al., 2017; Hoopmann et al., 2023), indicating a comparably low exposure to PCP-related chemicals in the investigated cohort. Nevertheless, up to seven PCPs were detected per urine sample and >50% of all samples contained four or more PCPs.

### Pesticides and pesticide metabolites

Pesticides are essential tools in conventional agriculture and are also frequently used in households and home gardens. Potential exposure routes include inhalation or dermal contact during (agricultural) pesticide application or intake of contaminated food or water. Frequent exposure to various pesticides during pregnancy has been reported in Bangladesh, and associations between prenatal pesticide exposure and impaired fetal development have been observed (Jaacks et al., 2019). In this study, we were able to detect the neonicotinoid imidacloprid (DF=40%, range: <LOQ-0.93 ng/mL), the neonicotinoid acetamiprid (ATMP; DF = 25%, range: <LOQ −0.082 ng/mL) and the organophosphate malathion (MAT; DF = 11%, range: 0.0057 ng/mL −0.26 ng/mL). Additionally, various pesticide metabolites, including p-nitrophenol (PNP; DF = 93%, range: <LOQ −15 ng/mL), the organophosphate metabolites DETP (DF = 83%, range: <LOQ −71 ng/mL) and DEDTP (DF = 34%, range: <LOQ −15 ng/mL), N,N-dimethylbenzamide (DBM; DF = 25%, range: <LOQ −0.70 ng/mL), 2,4-dichlorphenoxyacetic acid (2,4-D; DF = 16%, range: <LOQ −4.3 ng/mL) and 3-phenoxybenzoic acid (3-PBA; DF = 4.2%, range: <LOQ −9.0 ng/mL) were detected, highlighting frequent pesticide exposure of the studied population. In total, >99% of all urine samples contained at least one pesticide or pesticide metabolite. A high percentage (74%) of the samples contained three or more biomarkers for pesticide exposure, and up to eight pesticide biomarkers were detected per urine sample.

According to a recent systematic review, the most frequently used pesticides for vegetable production in Bangladesh are organophosphorus pesticides, pyrethroids, neonicotinoids, carbamates, and organochlorines, and approximately 30% of vegetable samples were found to contain pesticide residues (Khatun et al., 2023). Jaacks et al. (2019) also showed frequent pesticide exposure in a cohort of pregnant women from rural Bangladesh (n = 289) by analyzing pesticide metabolites in urine. They reported high detection frequencies for PNP (DF = 97.9%), a urinary biomarker for exposure to parathion and its metabolite methyl parathion. Jaacks et al. found that high exposure was associated with a higher risk for preterm delivery and smaller birth size of newborns (Jaacks et al., 2019). The high detection frequency of PNP found in the sample set (DF = 93%) underscores the frequent exposure to this class of pesticides in Bangladesh.

### Phytoestrogens and phytotoxins

Several phytochemicals were identified in the analyzed urine samples, including a range of phytoestrogenic compounds. While these compounds are generally associated with beneficial health effects, they also possess endocrine-disrupting properties (Nie et al., 2017). Isoflavones (e.g., daidzein, genistein, and related metabolites), prenylflavanoids (8-prenylnaringenin), lignans (e.g., matairesinol and gut-derived microbial metabolites enterodiol and enterolactone), stilbenoids (e.g., resveratrol), and coumestrol, as well as the toxic alkaloid metabolite jacobine N-oxide, were detected in urine samples in this study. Free forms of various phytoestrogenic compounds were ubiquitously detected with the highest detection frequencies for the gut-microbial lignan metabolites enterolactone (DF = 94%, range: <LOQ −320 ng/mL) and enterodiol (DF = 90%, range: <LOQ −12 ng/mL), followed by the isoflavones genistein (DF = 89%, range: <LOQ −26 ng/mL) and daidzein (DF = 84%, range: <LOQ −100 ng/mL). Notably, phytoestrogenic compounds are heavily conjugated before urinary excretion, and our data indicate that only a very small fraction is excreted in urine as the free parent compound. For instance, free enterodiol and enterolactone accounted for <1% of the total concentrations after deconjugation with *H.pomatia*.

The toxic alkaloid jacobine N-oxide was the only confirmed phytotoxin and was detected in 4.5% of samples at very low concentrations (<LOQ −0.42 ng/mL), made possible by its favorable ionization behavior. It is a metabolite of jacobine, a genotoxic pyrrolizidine alkaloid that is a potential contaminant in herbal infusions, tea, or honey (European Food Safety Authority (EFSA), 2016). Only a limited number of pyrrolizidine alkaloids were included in the assay for practical reasons.

### Plastic-related compounds

Humans are frequently exposed to chemicals leaching from plastic material, including compounds with known endocrine-disrupting properties, such as phthalates, and prenatal exposure can substantially impact fetal development and health (Almeida-Toledano et al., 2024). In this study, we detected six compounds related to plastic production, including phthalate plasticizers (MBP and MEHP), the non-phthalate plasticizer n-butylbenzenesulfonamide, and the bisphenols BPA, BPS, and BPF, which are used to produce various types of plastic products. Additionally, the plasticizers dibutylphthalate and benzyl-butyl phthalate were detected in most samples, but results are not reported herein due to comparably high concentrations in the extraction blanks. Highest detection frequencies were observed for N-butylbenzenesulfonamide (DF = 94%, range: <LOQ −5.9 ng/mL), followed by the dibutylphthalate metabolite MBP (DF = 62%, range: <LOQ −30 ng/mL), the DEHP metabolite MEHP (DF = 58%, range: <LOQ −14 ng/mL), BPS (DF = 38%, range: <LOQ −0.59 ng/mL), BPA (DF = 31%, range: <LOQ −2.9 ng/mL), and BPF, a substituent for the use of BPA in plastic production (DF = 7.4%, range: <LOQ −0.84 ng/mL). Biomarkers for exposure to plasticizers and related chemicals were ubiquitously detected in the investigated population, with >99% of samples containing at least one biomarker and >30% containing more than 3. The established HBM-GV for total concentration of urinary MBP is 190 ng/mL (Apel et al., 2023), and this value was not exceeded in our study when only free-form analyte concentrations were considered. Yet, our data indicate a high degree of conjugation in urine, with <1% of the analyte present in the free form (see **Table S15**). This is lower than reported in other studies, with 5%-10% free urinary MBP and large inter-individual variability (Zhu et al., 2016). Using an estimate of 1% free MBP, 9.6% of all samples exceeded HBM-GVs for urinary MBP, whereas with an estimate of 9.3% free MBP, only 0.7% of the samples exceeded HBM-GVs. Analogously, bisphenols are typically excreted as conjugates and only small fractions are accessible in their free form. For instance, we estimated free BPA accounting for <1% in this study, which is in line with Arbuckle et al. reporting approximately 1% free BPA in urine (Arbuckle Tye E. et al., 2015). Established HBM-GVs are based on total concentrations, which limits applicability to this dataset (Ougier et al., 2021). Using estimates of 1% free urinary BPA and 3% free urinary BPS (Khmiri et al., 2020), HBM-GV values were exceeded in 0.7% of the cases for BPA and in 13.6% of the cases for BPS (see **Table S7**). Notably, due to the high reported interindividual variability of free fractions, these estimates should be interpreted with caution given the high uncertainty.

### Co-exposure of xenobiotics and correlation with endogenous estrogen levels

To investigate the co-occurrence of xenobiotics and potential correlations with endogenous estrogen concentrations, spearman correlation analysis was performed. **Figure S7** shows a correlation heatmap for compounds with DF>50% (n = 24) based on specific density-normalized urinary concentrations, as well as data without applying any normalization. It is expected that urinary concentrations of functionally related analytes are correlated and speaks for the data quality. For instance, positive correlations were observed for urinary concentrations of the nicotine metabolites cotinine and t3OHCot (r = 0.95), the soybean-derived isoflavones daidzein and genistein (r = 0.80), and the gut microbial metabolites enterodiol and enterolactone (r = 0.69). Furthermore, urinary concentrations of the plasticizer n-butylbenzenesulfonamide were correlated with concentrations of 2-naphthol (r = 0.80) and ethylparaben (r = 0.67), and concentrations of propylparaben were correlated with those of methylparaben (r = 0.74) and ethylparaben (r = 0.37), indicating similar exposure routes. A negative correlation was observed between urinary concentrations of n-butlybenzensulfonamide and enterodiol (r = 0.33). Concentrations of the detected endogenous estrogens were highly inter-correlated with correlation coefficients r >0.5 for most analyte pairs. Moreover, a positive correlation with several xenobiotics with reported xenoestrogenic properties was observed. For instance, estradiol (E2) concentrations were correlated with enterodiol (r = 0.44), enterolactone (r = 0.50), 1-hydroxypyrene (r = 0.39), and genistein (r = 0.32).

### Limitations

In this study, we successfully applied a recently expanded next-generation biomonitoring method to a large number of urine samples from a cohort of pregnant women in rural Bangladesh for the first time. Yet, several limitations need to be noted:

First, only a single first-morning urine sample was collected in the first half of a pregnancy (i.e., <20 weeks gestation), and longitudinal variations during pregnancy were not assessed. Urinary concentrations can be used to monitor recent and chronic (i.e., continuous or repeatedly observed) exposure to xenobiotics; however, only limited information on previous exposure events can be derived. Therefore, analyzing a single sample can only provide snapshot data, and longitudinal studies with repeated urine collections could add valuable insights into the relationship between the exposome and health outcomes. Nevertheless, many exposure sources are closely linked to an individual’s lifestyle and behaviour patterns, such as dietary habits, living conditions, or environmental surroundings, and often remain stable over extended periods. For instance, recent work indicated surprisingly stable mycotoxin exposure during pregnancies in a US population, suggesting that exposure to food-borne contaminants can persist over long periods (Krausová et al., 2024).

Second, the presented dataset was acquired without applying enzymatic deconjugation as part of the sample preparation. In a recent study, substantial procedural backgrounds in enzyme mixtures used in typical deconjugation protocols, including sulfatase/glucuronidase solutions originating from *H.pomatia*, *E.coli,* and recombinant enzymes, was demonstrated. High concentrations of several natural and synthetic toxins were detected in these enzyme solutions, which is highly relevant to multi-class HBM methods. The results indicated a trade-off between deconjugation efficiency, coverage, and contamination regardless of the protocol used (Fareed et al., 2022). Analogously, the effect of enzymatic deconjugation using *H. pomatia* extracts was tested in the presented work. While this approach clearly increased concentrations of several compounds, substantial contamination with diverse natural and synthetic toxins was observed, including mycotoxins, phytoestrogens, phytotoxins, pesticides, and industrial chemicals. Consequently, a sample preparation protocol that omits analyte deconjugation was employed for the full sample set presented in this study, and the assay explicitly measures urinary concentrations of the free (unconjugated) form of the target analytes. Hence, for analytes that are primarily excreted as conjugates, total exposure is clearly underestimated in the presented data. To allow comparisons of reported urinary concentrations with other datasets and established HBM-GVs, deconjugation would be required. The absence of this step, therefore, limits the comparability of our dataset to reference concentrations or HBM-GVs, posing a challenge for toxicological interpretation of several analytes. We aimed to account for this limitation using estimates of the ratio of free form to total concentrations based on new experimental data and literature data where available. Notably, the free form accounts for only small fractions (e.g., 1-10%) of urinary concentrations for various xenobiotics, and metabolism and excretion may vary between individuals. Measuring only the free form results in lower detection frequencies because concentrations are lower than total concentrations after deconjugation, potentially leading to an underestimation of actual exposures.

Third, a clear distinction between active/intended intake and passive exposure to chemicals, such as antibiotics or stimulating substances like nicotine, remains challenging. A more detailed analysis of potential exposure routes would be beneficial.

Fourth, additional data analysis, including a detailed risk assessment that considers mixture effects, would be necessary to identify and delineate the cumulative effects of reported substances in real-life exposure scenarios. The evaluation of such effects, as well as the analysis of associations between exposure patterns and birth outcomes, is beyond the scope of this paper. Future work would also benefit from NTA/NTS using high-resolution mass spectrometry (HRMS) to maximize the chemical coverage and to identify additional exposures and potential metabolic responses, including the presence of phase II metabolites in urine samples.

## Conclusion

In total, 62 compounds were detected and (semi-) quantified in urine samples of pregnant women from Bangladesh participating in the MEMAPO cohort study. The detected analytes include relevant environmental and food-related contaminants such as pesticides, plasticizers, industrial chemicals, polyaromatic hydrocarbons, and mycotoxins. Several of the determined exposure classes (e.g., plasticizers, pesticides, industrial chemicals, nicotine metabolites, or PAH metabolites) were ubiquitously present in the studied sample set. Furthermore, antibiotics were detected in >60%, and mycoestrogens were detected >30% of the samples, indicating the need to monitor these analyte classes in routine HBM. Especially, urinary biomarkers for PAH exposure and nicotine intake were detected ubiuqoutously and at high concentrations. The availability of urinary concentrations, as well as normalized data based on specific density and urinary creatinine, provides a valuable resource for future data analysis. The presented data show widespread exposure of pregnant women from Bangladesh to diverse synthetic and natural toxicants and complements recently published HBM data on mycotoxin exposure in the same cohort. It will provide the basis for further investigations of the influence of the chemical exposome on pregnancy and birth outcomes in the future. Assessing such a wide range of biomarkers in pregnant women will set the foundation for investigating the combined effects of exposure to various chemicals on birth outcomes.

## Data Availability

All data produced in the present study are available upon reasonable request to the authors

## Declarations

### Ethical approval

The study adhered to the guidelines of the Declaration of Helsinki. The FAARM protocol received positive reviews from ethics committees in Bangladesh and Germany, with written informed consent obtained from all participants before data collection (Wendt et al., 2019). Additionally, the MEMAPO study was reviewed by ethics committees at Heidelberg University Medical Faculty in Germany as part of FAARM amendments and by the Institute of Health Economics at the University of Dhaka, Bangladesh (Reference: FWA00026031). MEMAPO participants were informed about the study’s objectives, the voluntary nature of participation, and the intended future analysis. Written informed consent was obtained from all participants before data collection. This research project has been conducted according to all relevant ethical guidelines and was approved by the ethics committee of the University of Vienna (no. 00157)

## Acknowledgments

Special thanks go to Amanda Wendt and Jillian Waid for their help and advice during data collection and for their help with the FAARM background data. We are also grateful to Maik Brune for his assistance with urine creatinine and density analysis and to Nurshad Ali, Abdul Kader, Rakibul Hassan, and Shafinaz Sobhan for their support with fieldwork. Finally, we thank the women enrolled in the MEMAPO study for their valuable time and participation.

The FAARM trial and the MEMAPO study were mainly funded by the German Federal Ministry of Education and Research (BMBF) (grant number: 01ER1201). Sabine Gabrysch received funding through a Recruiting Grant from Stiftung Charité, which supported part of the laboratory analysis.

The exposome analysis presented in this work was enabled by the Exposome Austria Research Infrastructure, the National Node of the ESFRI-EIRENE RI; the Austrian Federal Ministry of Education, Science and Research (project DigiOmics4AT); and the Austrian Federal Ministry for Climate Protection, Environment, Energy, Mobility, Innovation and Technology (BMK). This research was also funded in part by the Austrian Science Fund (FWF) [M 3217-N] via a Lise Meitner postdoctoral fellowship.

For the purpose of Open Access, the author has applied a CC BY public copyright license to any Author Accepted Manuscript version arising from this submission.

## Statement

During the preparation of this work, the authors used ChatGPT in order to refine the language in minor sections. After using this tool/service, the authors reviewed and edited the content as needed and take full responsibility for the content of the published article.

## References

Acevedo, J.M., Kahn, L.G., Pierce, K.A., Albergamo, V., Carrasco, A., Manuel, R.S.J., Singer Rosenberg, M., Trasande, L., 2025. Filling gaps in population estimates of phthalate exposure globally: A systematic review and meta-analysis of international biomonitoring data. Int. J. Hyg. Environ. Health 265, 114539. 10.1016/j.ijheh.2025.114539

Adams, K.J., Pratt, B., Bose, N., Dubois, L.G., St John-Williams, L., Perrott, K.M., Ky, K., Kapahi, P., Sharma, V., MacCoss, M.J., Moseley, M.A., Colton, C.A., MacLean, B.X., Schilling, B., Thompson, J.W., Alzheimer’s Disease Metabolomics Consortium, 2020. Skyline for Small Molecules: A Unifying Software Package for Quantitative Metabolomics. J. Proteome Res. 19, 1447–1458. 10.1021/acs.jproteome.9b00640

Almeida-Toledano, L., Navarro-Tapia, E., Sebastiani, G., Ferrero-Martínez, S., Ferrer-Aguilar, P., García-Algar, Ó., Andreu-Fernández, V., Gómez-Roig, M.D., 2024. Effect of prenatal phthalate exposure on fetal development and maternal/neonatal health consequences: A systematic review. Sci. Total Environ. 950, 175080. 10.1016/j.scitotenv.2024.175080

Angeles, L.F., Islam, S., Aldstadt, J., Saqeeb, K.N., Alam, M., Khan, M.A., Johura, F.-T., Ahmed, S.I., Aga, D.S., 2020. Retrospective suspect screening reveals previously ignored antibiotics, antifungal compounds, and metabolites in Bangladesh surface waters. Sci. Total Environ. 712, 136285. 10.1016/j.scitotenv.2019.136285

Apel, P., Angerer, J., Wilhelm, M., Kolossa-Gehring, M., 2017. New HBM values for emerging substances, inventory of reference and HBM values in force, and working principles of the German Human Biomonitoring Commission. Spec. Issue Hum. Biomonitoring 2016 220, 152–166. 10.1016/j.ijheh.2016.09.007

Apel, P., Lamkarkach, F., Lange, R., Sissoko, F., David, M., Rousselle, C., Schoeters, G., Kolossa-Gehring, M., 2023. Human biomonitoring guidance values (HBM-GVs) for priority substances under the HBM4EU initiative – New values derivation for deltamethrin and cyfluthrin and overall results. Int. J. Hyg. Environ. Health 248, 114097. 10.1016/j.ijheh.2022.114097

Arbuckle Tye E., Marro Leonora, Davis Karelyn, Fisher Mandy, Ayotte Pierre, Bélanger Patrick, Dumas Pierre, LeBlanc Alain, Bérubé René, Gaudreau Éric, Provencher Gilles, Faustman Elaine M., Vigoren Eric, Ettinger Adrienne S., Dellarco Michael, MacPherson Susan, Fraser William D., 2015. Exposure to Free and Conjugated Forms of Bisphenol A and Triclosan among Pregnant Women in the MIREC Cohort. Environ. Health Perspect. 123, 277–284. 10.1289/ehp.1408187

Ayeni, K.I., Berry, D., Wisgrill, L., Warth, B., Ezekiel, C.N., 2022. Early-life chemical exposome and gut microbiome development: African research perspectives within a global environmental health context. Trends Microbiol. 30, 1084–1100. 10.1016/j.tim.2022.05.008

Barnett-Itzhaki, Z., Esteban López, M., Puttaswamy, N., Berman, T., 2018. A review of human biomonitoring in selected Southeast Asian countries. Environ. Int. 116, 156–164. 10.1016/j.envint.2018.03.046

Bergkvist, C., Aune, M., Nilsson, I., Sandanger, T.M., Hamadani, J.D., Tofail, F., Oyvind-Odland, J., Kabir, I., Vahter, M., 2012. Occurrence and levels of organochlorine compounds in human breast milk in Bangladesh. Chemosphere 88, 784–790. 10.1016/j.chemosphere.2012.03.083

Calafat, A.M., 2012. The U.S. National Health and Nutrition Examination Survey and human exposure to environmental chemicals. Spec. Issue Berl. Int. Conf. Hum. Biomonitoring 215, 99–101. 10.1016/j.ijheh.2011.08.014

Chen, Y., Graziano, J.H., Parvez, F., Liu, M., Slavkovich, V., Kalra, T., Argos, M., Islam, T., Ahmed, A., Rakibuz-Zaman, M., Hasan, R., Sarwar, G., Levy, D., van Geen, A., Ahsan, H., 2011. Arsenic exposure from drinking water and mortality from cardiovascular disease in Bangladesh: prospective cohort study. BMJ 342, d2431. 10.1136/bmj.d2431

de Paula Nunes, E., Abou Dehn Pestana, B., Pereira, B.B., 2025. Human biomonitoring and environmental health: a critical review of global exposure patterns, methodological challenges and research gaps. J. Toxicol. Environ. Health Part B 1–19. 10.1080/10937404.2025.2529845

Esteban, M., Castaño, A., 2009. Non-invasive matrices in human biomonitoring: A review. Environ. Int. 35, 438–449. 10.1016/j.envint.2008.09.003

European Food Safety Authority (EFSA), 2016. Dietary exposure assessment to pyrrolizidine alkaloids in the European population. EFSA J. 14, e04572. 10.2903/j.efsa.2016.4572

Fareed, Y., Braun, D., Flasch, M., Globisch, D., Warth, B., 2022. A broad, exposome-type evaluation of xenobiotic phase II biotransformation in human biofluids by LC-MS/MS. Exposome 2, osac008. 10.1093/exposome/osac008

Forsyth, J.E., Weaver, K.L., Maher, K., Islam, M.S., Raqib, R., Rahman, M., Fendorf, S., Luby, S.P., 2019. Sources of Blood Lead Exposure in Rural Bangladesh. Environ. Sci. Technol. 53, 11429–11436. 10.1021/acs.est.9b00744

Garí, M., González-Quinteiro, Y., Bravo, N., Grimalt, J.O., 2018. Analysis of metabolites of organophosphate and pyrethroid pesticides in human urine from urban and agricultural populations (Catalonia and Galicia). Sci. Total Environ. 622–623, 526–533. 10.1016/j.scitotenv.2017.11.355

Gaudreau, É., Bérubé, R., Bienvenu, J.-F., Fleury, N., 2016. Stability issues in the determination of 19 urinary (free and conjugated) monohydroxy polycyclic aromatic hydrocarbons. Anal. Bioanal. Chem. 408, 4021–4033. 10.1007/s00216-016-9491-2

Geng, M., Liu, K., Huang, K., Zhu, Y., Ding, P., Zhang, J., Wang, B., Liu, W., Han, Y., Gao, H., Wang, S., Chen, G., Wu, X., Tao, F., 2020. Urinary antibiotic exposure across pregnancy from Chinese pregnant women and health risk assessment: Repeated measures analysis. Environ. Int. 145, 106164. 10.1016/j.envint.2020.106164

Gilles, L., Govarts, E., Rambaud, L., Vogel, N., Castaño, A., Esteban López, M., Rodriguez Martin, L., Koppen, G., Remy, S., Vrijheid, M., Montazeri, P., Birks, L., Sepai, O., Stewart, L., Fiddicke, U., Loots, I., Knudsen, L.E., Kolossa-Gehring, M., Schoeters, G., 2021. HBM4EU combines and harmonises human biomonitoring data across the EU, building on existing capacity – The HBM4EU survey. Int. J. Hyg. Environ. Health 237, 113809. 10.1016/j.ijheh.2021.113809

Gu, Y., Feuerstein, M.L., Lloyd, D.T., Patel, C.J., Johnson, C.H., Warth, B., 2025. Quantitative Exposomics Targeting over 200 Toxicants and Key Biomarkers at the Picomolar Level. Environ. Sci. Technol. 59, 21818–21829. 10.1021/acs.est.5c04458

Guerrini, C., Sunyer-Caldú, A., Gil-Solsona, R., Escribano, J., Vinaixa, M., Gago-Ferrero, P., Ramírez, N., 2024. Early-life chemical exposome: Comprehensive strategies for wide-scope screening of organic compounds. TrAC Trends Anal. Chem. 180, 117903. 10.1016/j.trac.2024.117903

Hartmann, C., Jamnik, T., Weiss, S., Göß, M., Fareed, Y., Satrapa, V., Braun, D., Flasch, M., Warth, B., Uhl, M., 2023. Results of the Austrian Children’s Biomonitoring Survey 2020 part A: Per- and polyfluorinated alkylated substances, bisphenols, parabens and other xenobiotics. Int. J. Hyg. Environ. Health 249, 114123. 10.1016/j.ijheh.2023.114123

Hoopmann, M., Murawski, A., Schümann, M., Göen, T., Apel, P., Vogel, N., Kolossa-Gehring, M., Röhl, C., 2023. A revised concept for deriving reference values for internal exposures to chemical substances and its application to population-representative biomonitoring data in German children and adolescents 2014–2017 (GerES V). Int. J. Hyg. Environ. Health 253, 114236. 10.1016/j.ijheh.2023.114236

Hossain, M.Z., Feuerstein, M.L., Gu, Y., Warth, B., 2024. Scaling up a targeted exposome LC-MS/MS biomonitoring method by incorporating veterinary drugs and pesticides. Anal. Bioanal. Chem. 416, 4369–4382. 10.1007/s00216-024-05374-x

Hossain, M.Z., Feuerstein, M.L., Warth, B., 2025. The role of residual (veterinary) antibiotics in chemical exposome analysis: Current progress and future perspectives. Compr. Rev. Food Sci. Food Saf. 24, e70105. 10.1111/1541-4337.70105

Hu, Y., Zhu, Q., Wang, Y., Liao, C., Jiang, G., 2022. A short review of human exposure to antibiotics based on urinary biomonitoring. Sci. Total Environ. 830, 154775. 10.1016/j.scitotenv.2022.154775

Jaacks, L.M., Diao, N., Calafat, A.M., Ospina, M., Mazumdar, M., Ibne Hasan, M.O.S., Wright, R., Quamruzzaman, Q., Christiani, D.C., 2019. Association of prenatal pesticide exposures with adverse pregnancy outcomes and stunting in rural Bangladesh. Environ. Int. 133, 105243. 10.1016/j.envint.2019.105243

Jagani, R., Pulivarthi, D., Patel, D., Wright, R.J., Wright, R.O., Arora, M., Wolff, M.S., Andra, S.S., 2022. Validated single urinary assay designed for exposomic multi-class biomarkers of common environmental exposures. Anal. Bioanal. Chem. 414, 5943–5966. 10.1007/s00216-022-04159-4

Jamnik, T., Flasch, M., Braun, D., Fareed, Y., Wasinger, D., Seki, D., Berry, D., Berger, A., Wisgrill, L., Warth, B., 2022. Next-generation biomonitoring of the early-life chemical exposome in neonatal and infant development. Nat. Commun. 13, 2653. 10.1038/s41467-022-30204-y

Joksić, A.Š., Tratnik, J.S., Mazej, D., Kocman, D., Stajnko, A., Eržen, I., Horvat, M., 2022. Polycyclic aromatic hydrocarbons (PAHs) in men and lactating women in Slovenia: Results of the first national human biomonitoring. Int. J. Hyg. Environ. Health 241, 113943. 10.1016/j.ijheh.2022.113943

Khatun, P., Islam, A., Sachi, S., Islam, Md.Z., Islam, P., 2023. Pesticides in vegetable production in Bangladesh: A systemic review of contamination levels and associated health risks in the last decade. Toxicol. Rep. 11, 199–211. 10.1016/j.toxrep.2023.09.003

Khmiri, I., Côté, J., Mantha, M., Khemiri, R., Lacroix, M., Gely, C., Toutain, P.-L., Picard-Hagen, N., Gayrard, V., Bouchard, M., 2020. Toxicokinetics of bisphenol-S and its glucuronide in plasma and urine following oral and dermal exposure in volunteers for the interpretation of biomonitoring data. Environ. Int. 138, 105644. 10.1016/j.envint.2020.105644

Kibria, G., Hossain, M.M., Mallick, D., Lau, T.C., Wu, R., 2016. Trace/heavy metal pollution monitoring in estuary and coastal area of Bay of Bengal, Bangladesh and implicated impacts. Mar. Pollut. Bull. 105, 393–402. 10.1016/j.marpolbul.2016.02.021

Krausová, M., Ayeni, K.I., Gu, Y., Borutzki, Y., O’Bryan, J., Perley, L., Silasi, M., Wisgrill, L., Johnson, C.H., Warth, B., 2024. Longitudinal biomonitoring of mycotoxin exposure during pregnancy in the Yale pregnancy Outcome Prediction study. Environ. Int. 109081. 10.1016/j.envint.2024.109081

Krausová, M., Braun, D., Buerki-Thurnherr, T., Gundacker, C., Schernhammer, E., Wisgrill, L., Warth, B., 2023. Understanding the Chemical Exposome During Fetal Development and Early Childhood: A Review. Annu. Rev. Pharmacol. Toxicol. 10.1146/annurev-pharmtox-051922-113350

Kyei, N.N.A., Boakye, D., Gabrysch, S., 2020. Maternal mycotoxin exposure and adverse pregnancy outcomes: a systematic review. Mycotoxin Res. 36, 243–255. 10.1007/s12550-019-00384-6

Kyei, N.N.A., Cramer, B., Humpf, H.-U., Degen, G.H., Ali, N., Gabrysch, S., 2022. Assessment of multiple mycotoxin exposure and its association with food consumption: a human biomonitoring study in a pregnant cohort in rural Bangladesh. Arch. Toxicol. 96, 2123–2138. 10.1007/s00204-022-03288-0

Kyei, N.N.A., Waid, J.L., Ali, N., Cramer, B., Humpf, H.-U., Gabrysch, S., 2023. Maternal exposure to multiple mycotoxins and adverse pregnancy outcomes: a prospective cohort study in rural Bangladesh. Arch. Toxicol. 97, 1795–1812. 10.1007/s00204-023-03491-7

Lai, Y., Koelmel, J.P., Walker, D.I., Price, E.J., Papazian, S., Manz, K.E., Castilla-Fernández, D., Bowden, J.A., Nikiforov, V., David, A., Bessonneau, V., Amer, B., Seethapathy, S., Hu, X., Lin, E.Z., Jbebli, A., McNeil, B.R., Barupal, D., Cerasa, M., Xie, H., Kalia, V., Nandakumar, R., Singh, R., Tian, Z., Gao, P., Zhao, Y., Froment, J., Rostkowski, P., Dubey, S., Coufalíková, K., Seličová, H., Hecht, H., Liu, S., Udhani, H.H., Restituito, S., Tchou-Wong, K.-M., Lu, K., Martin, J.W., Warth, B., Godri Pollitt, K.J., Klánová, J., Fiehn, O., Metz, T.O., Pennell, K.D., Jones, D.P., Miller, G.W., 2024. High-Resolution Mass Spectrometry for Human Exposomics: Expanding Chemical Space Coverage. Environ. Sci. Technol. 58, 12784–12822. 10.1021/acs.est.4c01156

Lerbech, A.M., Opintan, J.A., Bekoe, S.O., Ahiabu, M.-A., Tersbøl, B.P., Hansen, M., Brightson, K.T.C., Ametepeh, S., Frimodt-Møller, N., Styrishave, B., 2014. Antibiotic Exposure in a Low-Income Country: Screening Urine Samples for Presence of Antibiotics and Antibiotic Resistance in Coagulase Negative Staphylococcal Contaminants. PLOS ONE 9, e113055. 10.1371/journal.pone.0113055

McAdam, K.G., Faizi, A., Kimpton, H., Porter, A., Rodu, B., 2013. Polycyclic aromatic hydrocarbons in US and Swedish smokeless tobacco products. Chem. Cent. J. 7, 151. 10.1186/1752-153X-7-151

Miller, G.W., 2024. Exposomics: perfection not required. Exposome 4, osae006. 10.1093/exposome/osae006

Murawski, A., Roth, A., Schwedler, G., Schmied-Tobies, M.I.H., Rucic, E., Pluym, N., Scherer, M., Scherer, G., Conrad, A., Kolossa-Gehring, M., 2020. Polycyclic aromatic hydrocarbons (PAH) in urine of children and adolescents in Germany – human biomonitoring results of the German Environmental Survey 2014–2017 (GerES V). Int. J. Hyg. Environ. Health 226, 113491. 10.1016/j.ijheh.2020.113491

Nie, Q., Xing, M., Hu, J., Hu, X., Nie, S., Xie, M., 2017. Metabolism and health effects of phyto-estrogens. Crit. Rev. Food Sci. Nutr. 57, 2432–2454. 10.1080/10408398.2015.1077194

Nowak, K., Ratajczak–Wrona, W., Górska, M., Jabłońska, E., 2018. Parabens and their effects on the endocrine system. Mol. Cell. Endocrinol. 474, 238–251. 10.1016/j.mce.2018.03.014

Ougier, E., Zeman, F., Antignac, J.-P., Rousselle, C., Lange, R., Kolossa-Gehring, M., Apel, P., 2021. Human biomonitoring initiative (HBM4EU): Human biomonitoring guidance values (HBM-GVs) derived for bisphenol A. Environ. Int. 154, 106563. 10.1016/j.envint.2021.106563

Pacyga, D.C., Talge, N.M., Gardiner, J.C., Calafat, A.M., Schantz, S.L., Strakovsky, R.S., 2022. Maternal diet quality moderates associations between parabens and birth outcomes. Environ. Res. 214, 114078. 10.1016/j.envres.2022.114078

Parvin, F., Haque, M.M., Tareq, S.M., 2022. Recent status of water quality in Bangladesh: A systematic review, meta-analysis and health risk assessment. Environ. Chall. 6, 100416. 10.1016/j.envc.2021.100416

Preindl, K., Braun, D., Aichinger, G., Sieri, S., Fang, M., Marko, D., Warth, B., 2019. A Generic Liquid Chromatography−Tandem Mass Spectrometry Exposome Method for the Determination of Xenoestrogens in Biological Matrices. Anal. Chem. 91, 11334–11342. 10.1021/acs.analchem.9b02446

Rasel, S.M., Siddique, A.B., Nayon, Md.F.S., Suzon, M.S.M., Amin, S., Mim, S.S., Hossain, Md.S., 2024. Assessment of the association between health problems and cooking fuel type, and barriers towards clean cooking among rural household people in Bangladesh. BMC Public Health 24, 512. 10.1186/s12889-024-17971-7

Ringbeck, B., Belov, V.N., Schmidtkunz, C., Küpper, K., Gries, W., Weiss, T., Brüning, T., Hayen, H., Bury, D., Leng, G., Koch, H.M., 2021. Human Metabolism and Urinary Excretion Kinetics of Nonylphenol in Three Volunteers after a Single Oral Dose. Chem. Res. Toxicol. 34, 2392–2403. 10.1021/acs.chemrestox.1c00301

Wendt, A.S., Sparling, T.M., Waid, J.L., Mueller, A.A., Gabrysch, S., 2019. Food and Agricultural Approaches to Reducing Malnutrition (FAARM): protocol for a cluster-randomised controlled trial to evaluate the impact of a Homestead Food Production programme on undernutrition in rural Bangladesh. BMJ Open 9, e031037. 10.1136/bmjopen-2019-031037

Wild, C.P., 2012. The exposome: from concept to utility. Int. J. Epidemiol. 41, 24–32. 10.1093/ije/dyr236

Wild, C.P., 2005. Complementing the Genome with an “Exposome”: The Outstanding Challenge of Environmental Exposure Measurement in Molecular Epidemiology. Cancer Epidemiol. Biomarkers Prev. 14, 1847–1850. 10.1158/1055-9965.EPI-05-0456

Wild, C.P., Gong, Y.Y., 2010. Mycotoxins and human disease: a largely ignored global health issue. Carcinogenesis 31, 71–82. 10.1093/carcin/bgp264

Zhu, Y., Liu, K., Zhang, J., Liu, X., Yang, L., Wei, R., Wang, S., Zhang, D., Xie, S., Tao, F., 2020. Antibiotic body burden of elderly Chinese population and health risk assessment: A human biomonitoring-based study. Environ. Pollut. 256, 113311. 10.1016/j.envpol.2019.113311

Zhu, Y., Wan, Y., Li, Y., Zhang, B., Zhou, A., Cai, Z., Qian, Z., Zhang, C., Huo, W., Huang, K., Hu, J., Cheng, L., Chang, H., Huang, Z., Xu, B., Xia, W., Xu, S., 2016. Free and total urinary phthalate metabolite concentrations among pregnant women from the Healthy Baby Cohort (HBC), China. Environ. Int. 88, 67–73. 10.1016/j.envint.2015.12.004

