## Supplementary material for "Next-generation biomonitoring in a cohort of pregnant women from rural Bangladesh": Supplemenatry Information - Text and Figures

### Description of the *Food and Agricultural Approaches to Reducing Malnutrition* (FAARM) and *Maternal Exposure to Mycotoxins and Adverse Pregnancy Outcomes* (MEMAPO) study populations:

The *Food and Agricultural Approaches to Reducing Malnutrition* (FAARM) study was a 1:1 parallel-arm, cluster-randomized controlled trial conducted in Nabiganj and Baniachong sub-districts of Habiganj District, Sylhet Division, Bangladesh (ClinicalTrials.gov ID: NCT02505711). The FAARM trial (2015-2020) included 2,705 young married women in 96 settlements (geographic clusters) who reported being 30 years or younger, having access to at least 40 m² of land, and declared an interest in gardening. The trial assessed the impact of a homestead food production program, implemented by the nonprofit Helen Keller International, on maternal and child undernutrition. Further details on the FAARM trial design are available in the published study protocol (Wendt et al., 2019). As an extension of FAARM, the *Maternal Exposure to Mycotoxins and Adverse Pregnancy Outcomes* (MEMAPO) study, a nested prospective cohort study, was conducted from 2018 to 2020. MEMAPO followed 439 pregnant FAARM participants from early pregnancy to investigate the effects of maternal mycotoxin exposure on adverse pregnancy outcomes (Kyei et al., 2023). Between July 2018 and November 2019, 439 pregnant FAARM participants with a known last menstrual period and a gestational age of less than 20 weeks were enrolled in the MEMAPO cohort. As part of the MEMAPO data collection, about 10 mL of first-morning urine samples, before consuming food and water, were collected in a sterile disposable container from each study participant. The urine samples were intended for mycotoxin and other environmental exposure analyses. They were transported to the project field laboratory in a cold box, aliquoted into 2 mL safe-seal tubes, and stored at -20°C on the same day of urine pick-up. All samples were subsequently sent to Germany on dry ice (transport time < 24 hours) and stored at -70°C at Heidelberg University. Sample aliquots for chemical exposome analysis were later transported on dry ice to the University of Vienna, Austria (transport time < 6 hours) and stored at -80°C until biomarker analysis.

### Urine creatinine and urine density analysis:

Routine urine analyses were conducted at the accredited central laboratory of Heidelberg University Hospital. Specific gravity and other urine status parameters were measured using multi-parameter test strips (#10634643, Siemens Healthcare GmbH) on a Siemens CLINITEK Novus® analyzer, following the manufacturer's guidelines. Urine creatinine levels were determined using a Siemens ADVIA XPT chemistry analyzer (Siemens kit #03039070).

### Supplementary data:

#### Stability of internal standards

Peak areas (area under curve, AUC) and retention times for 10 selected internal standards (ISTDs) were tracked in 450 sample injections. Relative standard deviation (RSD; %) was used as a metric to investigate the stability of the LC-MS/MS assay. ISTD data was investigated and compounds were evaluated only if no interference was observed and if signal intensity was sufficient. The internal standard mixture was spiked into all samples and quality controls (QCs) before extraction, allowing to track the variability of the entire workflow including sample preparation, LC separation and MS/MS acquisition over >700 injections (~10 days total acquisition time). Results can be reviewed in **Table S5** and are summarized in **Figure S1** with panel a) showing histograms for determined retention times (min) and panel b) showing a histogram representation for AUCs. Example MRM-chromatograms for ^13^C_12_ bisphenol A (BPA) are presented in **Figure S2**. All 10 selected ISTDs showed stable retention times (RTs) and acceptable RSDs for peak areas with highest RSD observed for florfenicol-D3 (RSD=22.5%).

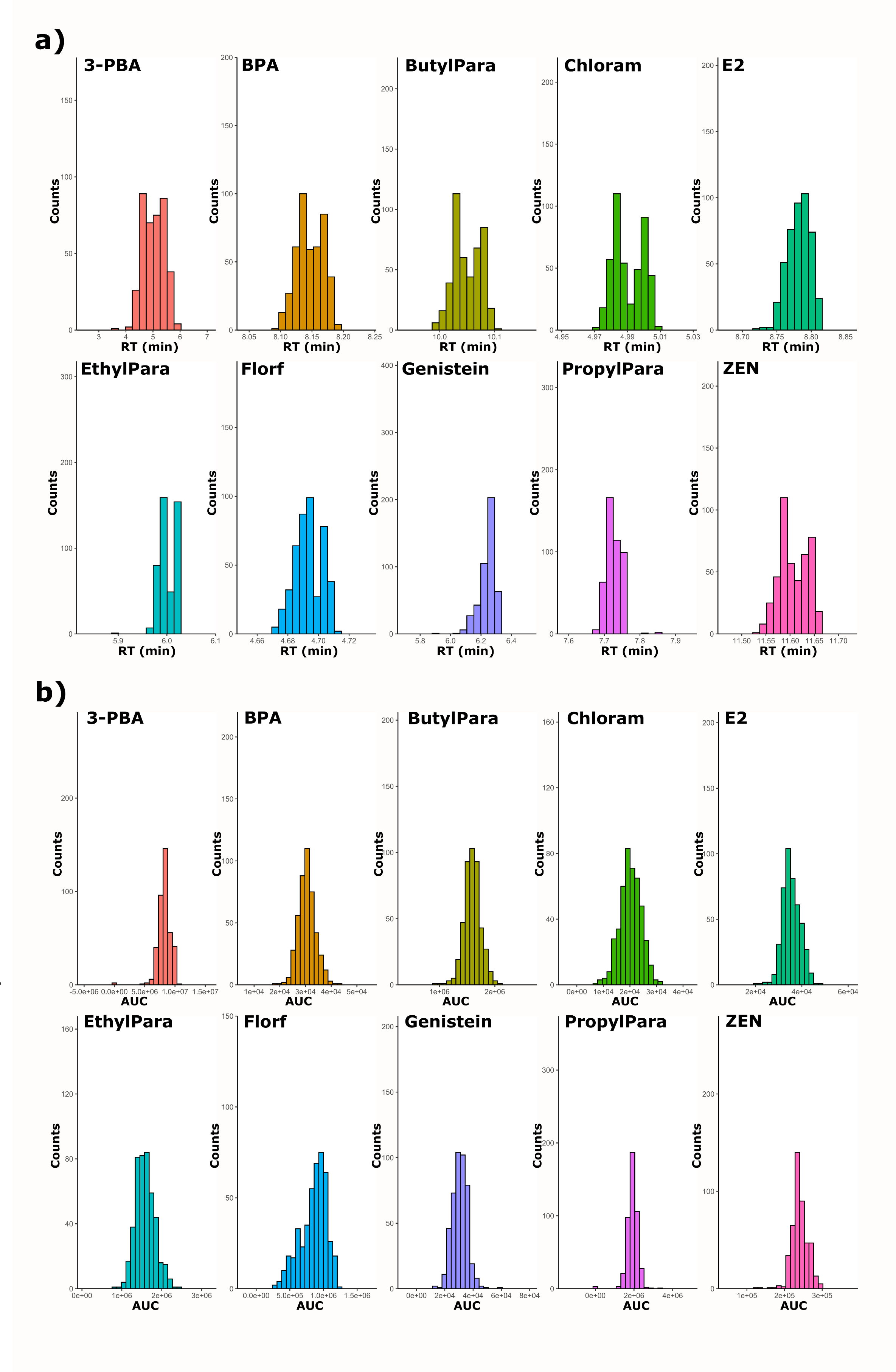

**Figure S1:** a) Histograms of retention times (RTs) of ten selected internal standards (ISTDs) in all sample injections and b) histograms of peak areas (area under curve; AUC) of ISTDs in all sample injections (n=450).

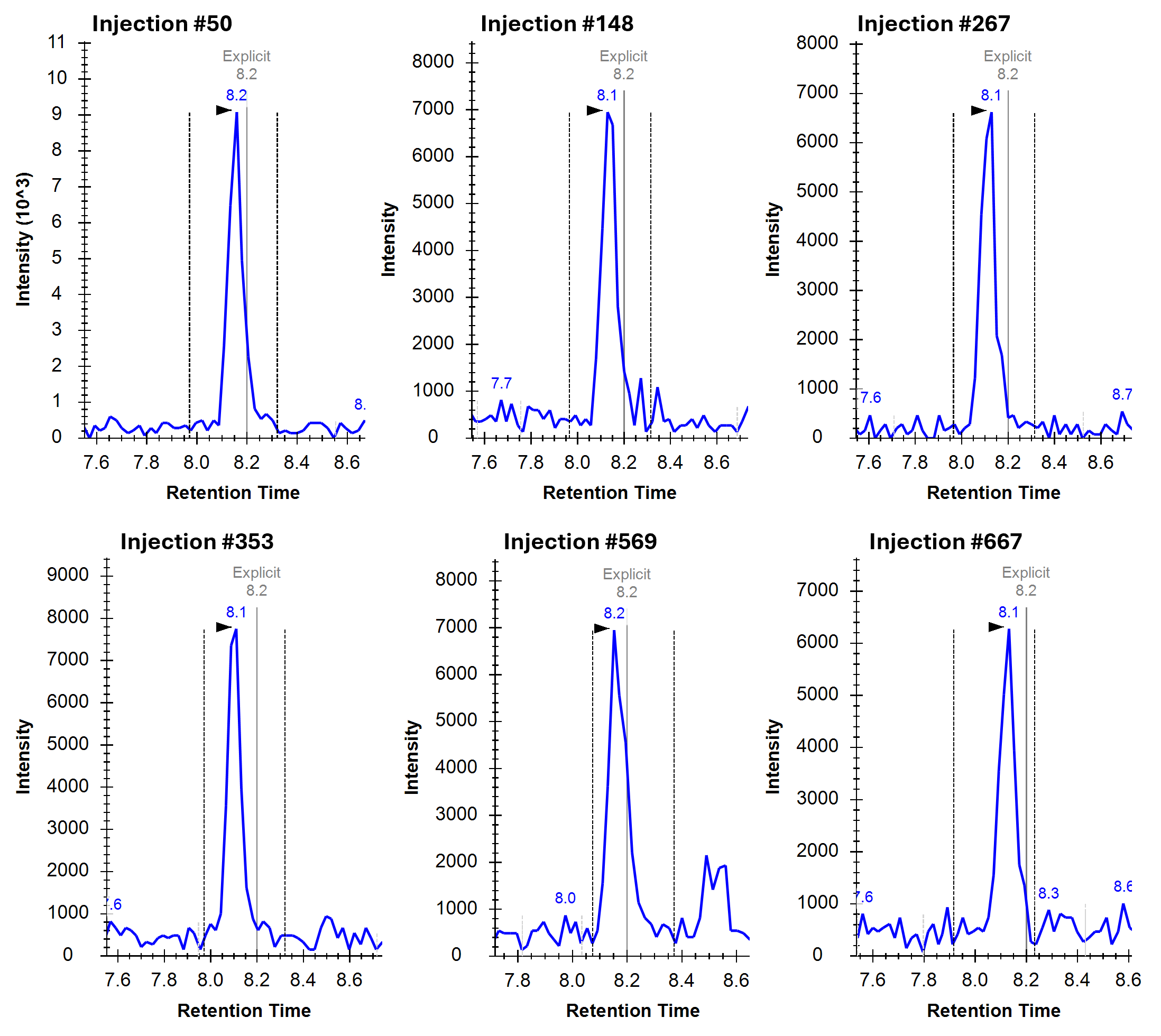

**Figure S2:** MRM-chromatograms of ^13^C_12_ bisphenol A (BPA) for samples with injections number 50, 148, 267, 353, 569, and 667.

#### Apparent recovery and blank correction

The apparent recovery was determined by measuring non-spiked and spiked urine samples in triplicates and was used to correct matrix effects and analyte recovery. The average apparent recovery for the 62 detected compounds was 84% ± 32% (median = 91%, min. = 17%, max. = 182%). Data for analytes with apparent recoveries between 60% and 140% were classified as quantitative data, whereas results for compounds outside of this range were reported as semiquantitative. A summary of the analytical figures of merits (apparent recovery, detection limit and quantification limit), as well as of the data quality (quantitative, semiquantitative, not detected, not reported) is given in **Table S6**.

Reported urinary concentrations *(c_rep_)* were corrected for apparent recovery and mean extraction blanks using the following formula:

(1) $c_{rep.}= \frac{100\cdot c_{m}}{RE(\%)}-c_{blank}$

with mean blank concentration ($c_{blank}$), measured concentration ($c_{m}$) based on calibration curves in neat solvent, and apparent recovery *RE.*

#### Blinded samples as quality control samples

Samples of two participants were additionally used as blinded quality control samples to investigate robustness of quantitative results. For this purpose, two aliquots of each sample were extracted and analyzed in duplicates and concentrations > LOQ were compared. 11 analytes could be quantified in both replicates of individual A, whereas 17 compounds were quantified in both replicates of individual B and RSDs were 13% ± 12% for sample A and 12% ± 7% for sample B. In total, 18 compounds were detected in the duplicate samples and a maximum RSD of 37% was observed for the plasticizer n-butylbenzene sulfonamide in sample A.

#### Spiked quality control samples

A pooled urine sample was spiked (calibration level 30, see **Table S3**) and repeatedly injected throughout the measurement sequence. **Figure S3** shows MRM-chromatograms and peak areas of spiked QC samples for five selected compounds (cotinine, propylparaben, amoxicillin, trimethoprim, benzophenone 1) and demonstrates excellent repeatability throughout the analytical sequence.

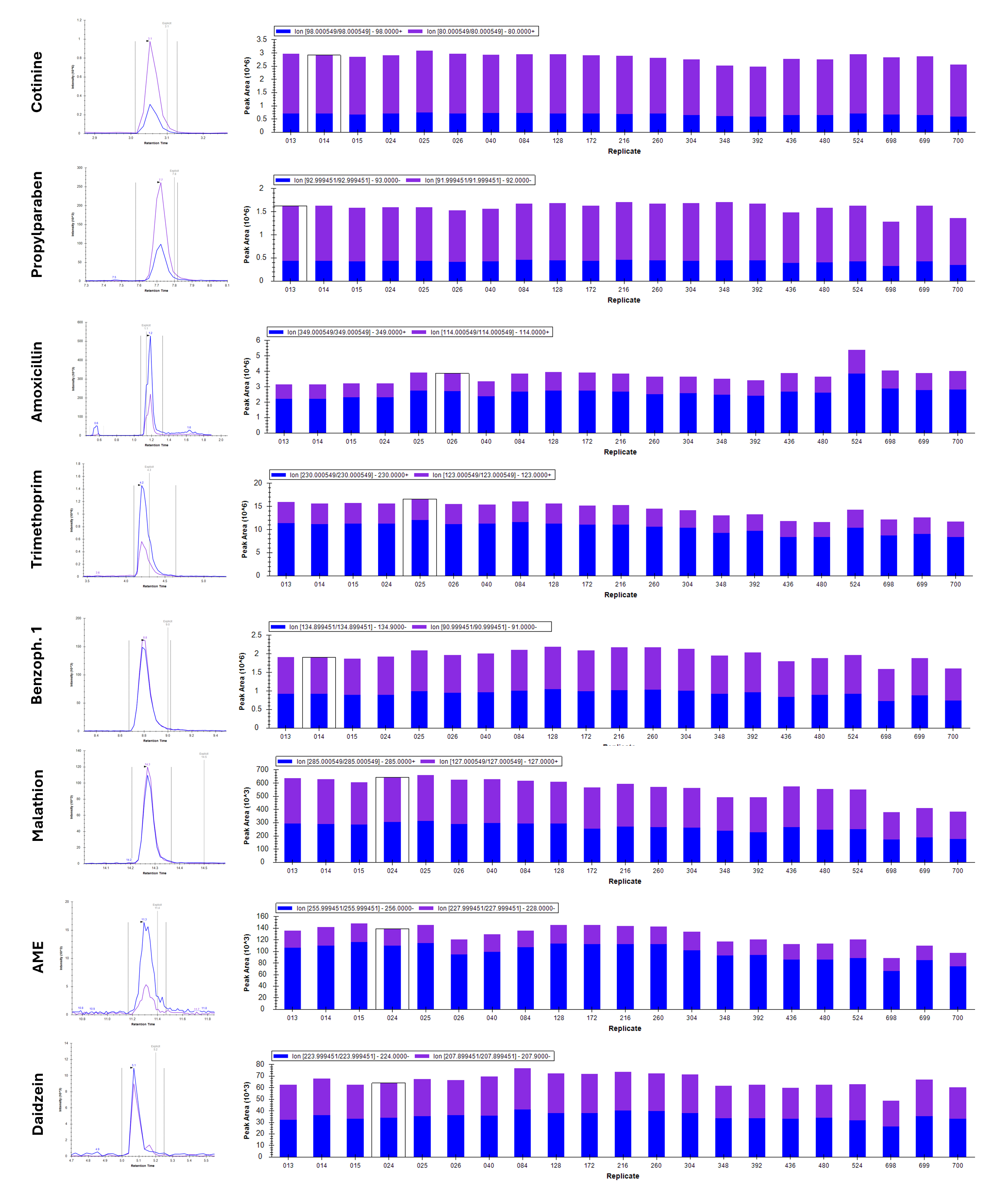

**Figure S3:** Example MRM-chromatograms and bar plots of peak areas for selected detected compounds in spiked quality control (QC) samples (n=21): cotinine, propylparaben, amoxicillin, trimethoprim, and benzophenone 1. All compounds were quantified in real-life samples during this study and two transitions per compound are shown (blue and purple lines or bars).

#### Detected analytes

In total, 62 analytes were detected in at least one urine sample measured in this study. **Figure S4** shows MRM-chromatograms for selected analytes at low concentrations, high concentrations, in a spiked sample as well as a procedural blank sample. **Figure S5** shows MRM-chromatograms for examples of diverse detected analytes at higher concentration levels. **Figure S6** shows the distribution of the number of detected compounds per sample resolved by compound class for antibiotics/veterinary drugs, personal care products, pesticides and pesticide metabolites, plastic related compounds as well as for mycotoxins and phytoestrogens.

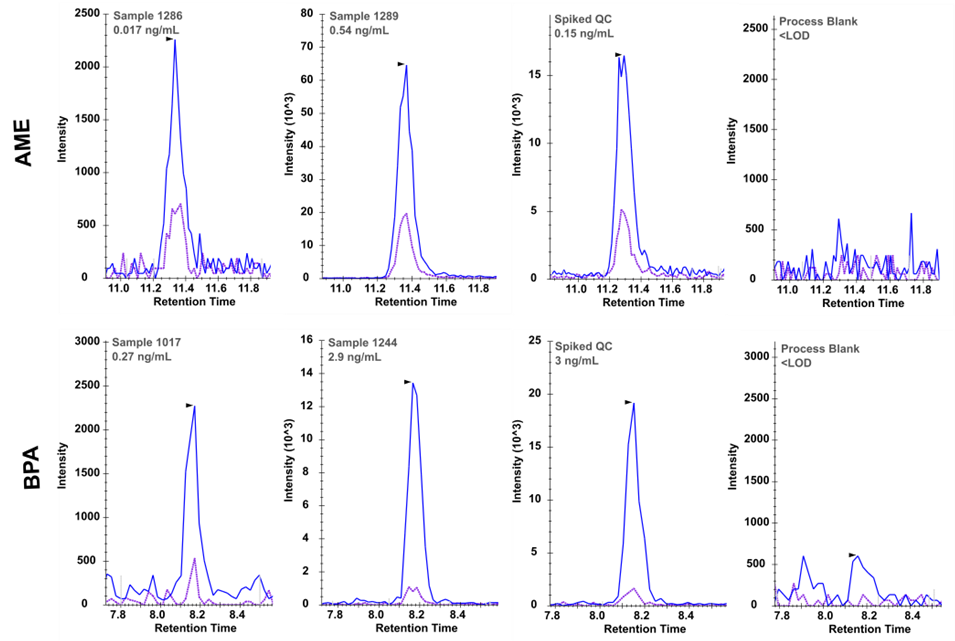

**Figure S 4:** Example MRM-chromatogram of a sample with low concentration, high concentration, a spiked QC sample and the process blanks for the mycotoxin alternariol monomethyl ether (AME) and bisphenol A (BPA). Data shown for two transitions per compound (represented as blue and purple lines).

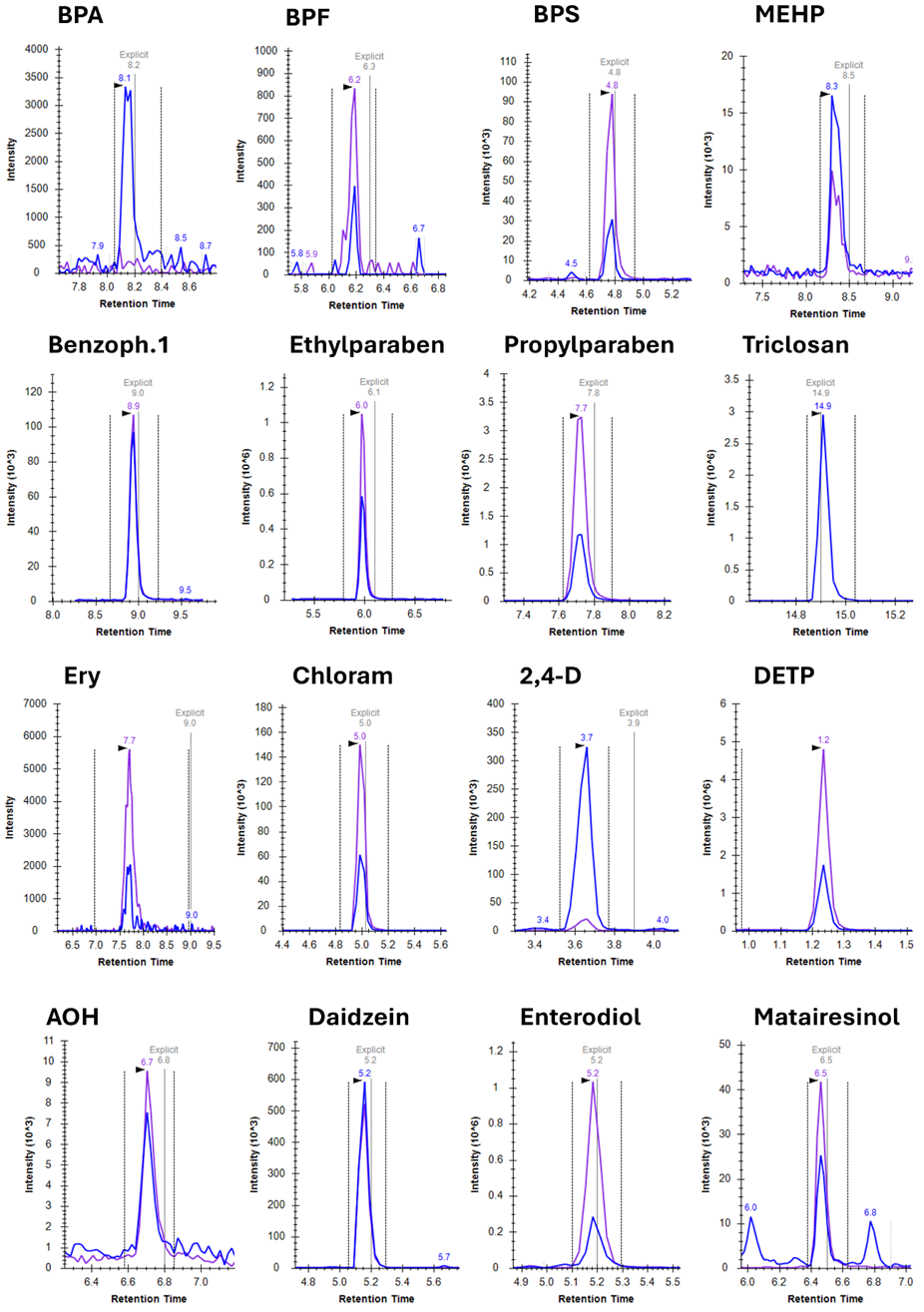

**Figure S5**: Example MRM-chromatograms for a subset of detected compounds in urine samples illustrating the wide chemical coverage of the used HBM assay. Detected analytes included plastic related compounds (e.g., bisphenol A), personal care products (e.g., propylparaben), antibiotics (e.g., erythromycin; Ery), pesticides (e.g., DETP), mycotoxins (e.g., alternariol; AOH), and phytoestrogens (e.g., daidzein). Data shown for two transitions per compound is shown (represented in as blue and purple lines).

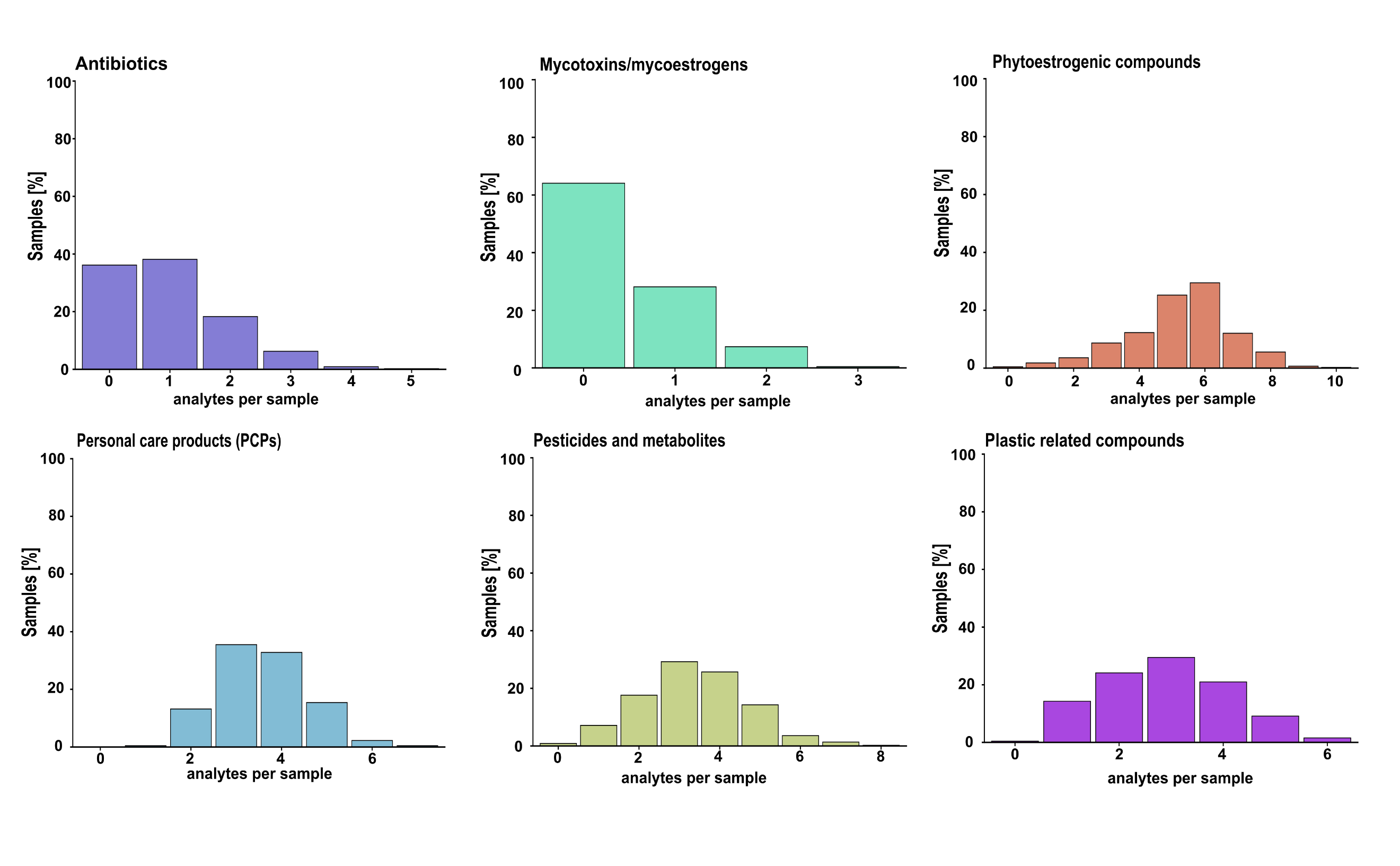

**Figure S6:** Number of detected analytes per sample per compound class as histograms for antibiotics, mycotoxins/mycoestrogens, phytoestrogens, personal care products (PCPs), pesticides and pesticide metabolites, and plastic related compounds

#### Correlations between detected compounds

Spearman correlation coefficients were used to investigate correlations between detected analytes, and p-values were calculated using the corrplot package in R. **Figure** **S7** shows a correlation heatmap for all compounds with detection frequencies (DF) >50% (n=24).

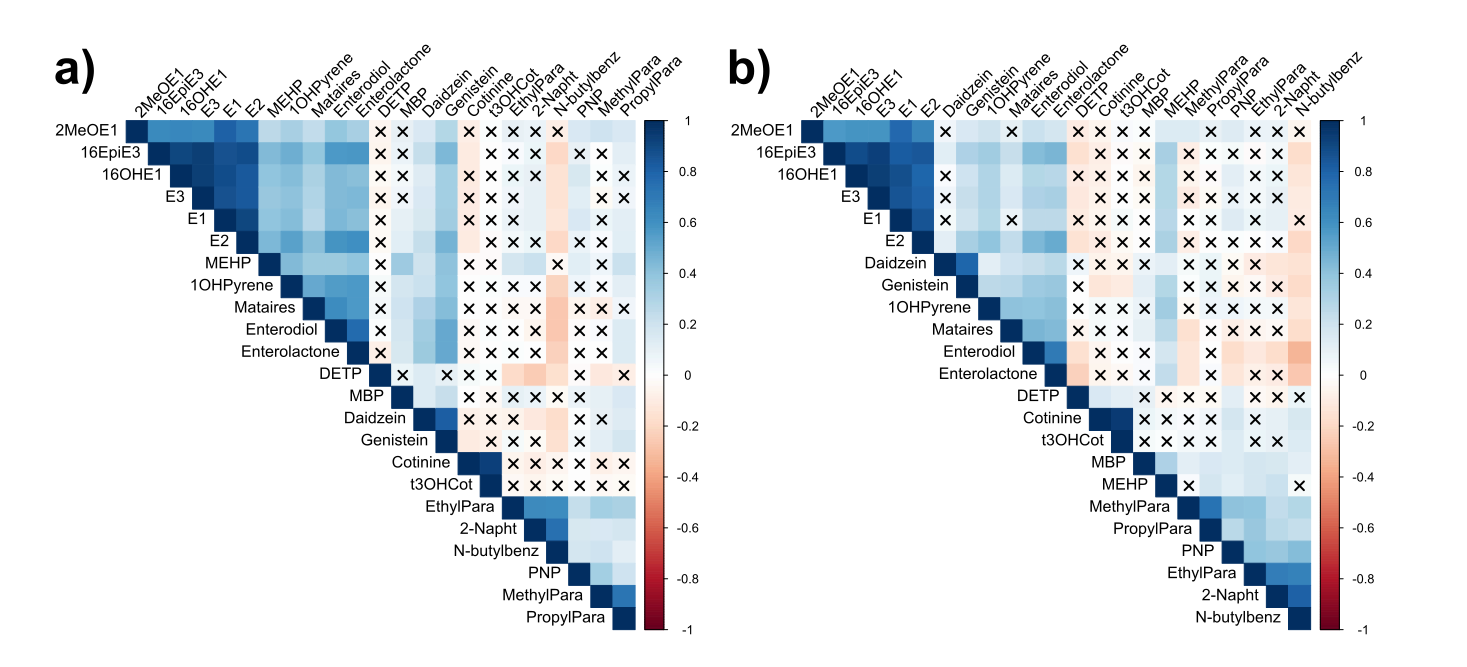

**Figure S7**: a) Spearman correlation heatmap of all 24 xenobiotics and endogenous estrogens with DF>50% in urine samples (n=446) and b) after using specific gravity normalized urinary concentrations. Concentrations <LOD were imputed with 0.5·LOD and concentrations <LOQ were imputed with 0.5·LOQ. FDR-corrected p-values were used to test significance of correlations with p-values >0.05 crossed out.

#### Enzymatic deconjugation

Enzymatic deconjugation (i.e., glucuronidase/sulfatase treatment) was performed for a subset of samples using one of our previously published protocols (Fareed et al., 2022; Hossain et al., 2024). In short, 100 µL of urine was treated with 100 µM β-glucuronidase/sulfatase enzyme solution from *Helix pomatia* in 2.5 M NH_4_Ac buffer adjusted to pH 5.5. The enzymes were prepared, samples were incubated for 16 h at 400 rpm and 37°C on a heater/shaker and samples were extracted with the same protocol as described for urine samples and with a final reconstitution volume of 100 µL. A comparison of background concentrations determined in procedural blanks with and without using β-glucuronidase/sulfatase treatment is provided in **Table S16**.

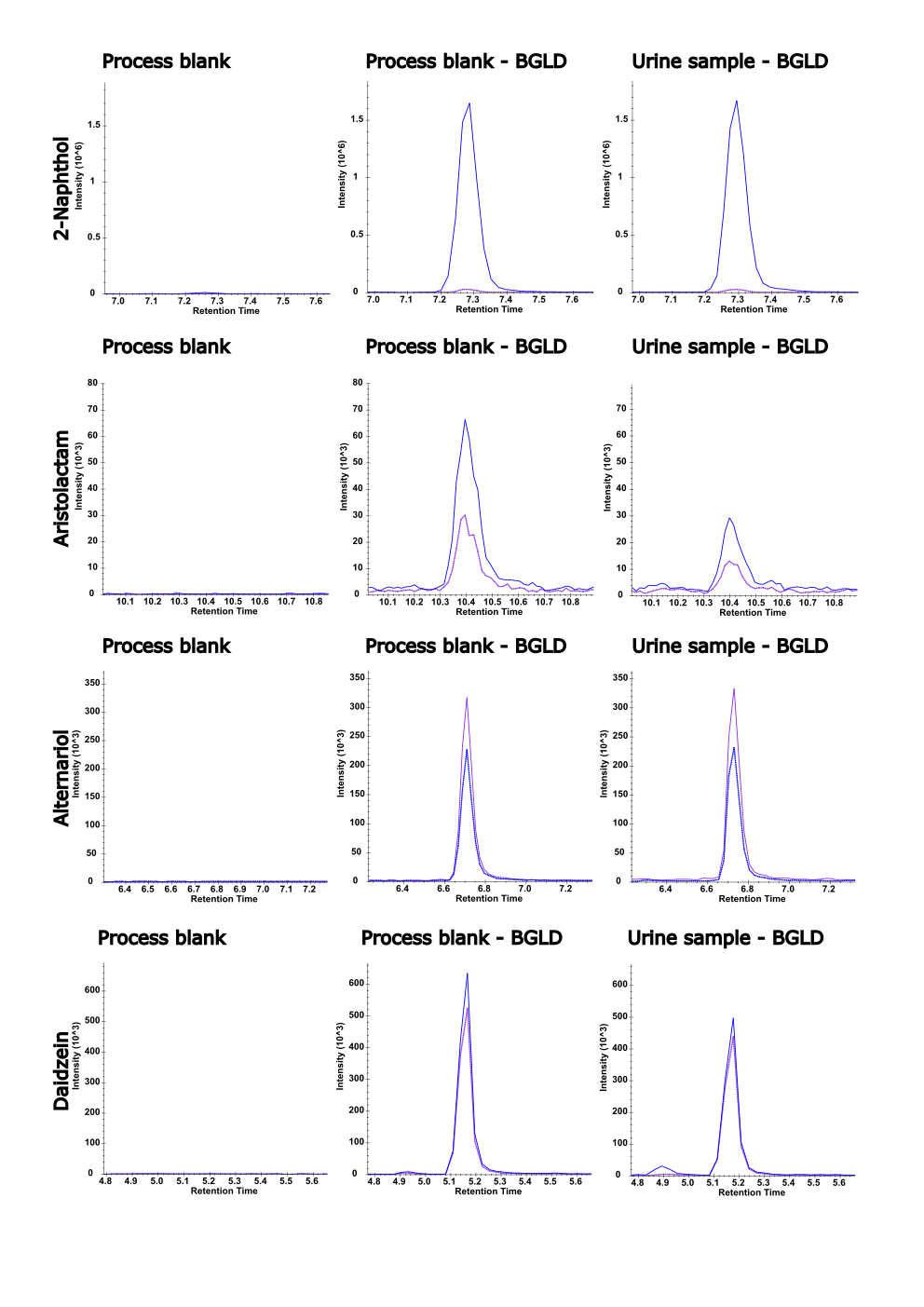

**Figure S8**: Examples for high contamination of the used HHH. pomatia enzyme solution. Panels show example chromatograms for process blanks of the extraction method (no deconjugation applied), process blank after deconjugation (BGLD) and concentrations in samples after conjugation. Data are presented for 2-naphthol, the plant alkaloid aristolactam, the mycotoxin alternariol and the phytoestrogen daidzein.

### References:

Fareed, Y., Braun, D., Flasch, M., Globisch, D., Warth, B., 2022. A broad, exposome-type evaluation of xenobiotic phase II biotransformation in human biofluids by LC-MS/MS. Exposome 2, osac008. https://doi.org/10.1093/exposome/osac008

Hossain, M.Z., Feuerstein, M.L., Gu, Y., Warth, B., 2024. Scaling up a targeted exposome LC-MS/MS biomonitoring method by incorporating veterinary drugs and pesticides. Analytical and Bioanalytical Chemistry 416, 4369–4382. https://doi.org/10.1007/s00216-024-05374-x

Kyei, N.N.A., Waid, J.L., Ali, N., Cramer, B., Humpf, H.-U., Gabrysch, S., 2023. Maternal exposure to multiple mycotoxins and adverse pregnancy outcomes: a prospective cohort study in rural Bangladesh. Archives of Toxicology 97, 1795–1812. https://doi.org/10.1007/s00204-023-03491-7

Wendt, A.S., Sparling, T.M., Waid, J.L., Mueller, A.A., Gabrysch, S., 2019. Food and Agricultural Approaches to Reducing Malnutrition (FAARM): protocol for a cluster-randomised controlled trial to evaluate the impact of a Homestead Food Production programme on undernutrition in rural Bangladesh. BMJ Open 9, e031037. https://doi.org/10.1136/bmjopen-2019-031037
